# Prevalence and Predictors of Pain and Fatigue in Pediatric Acute-Onset Neuropsychiatric Syndrome (PANS)

**DOI:** 10.64898/2026.10.02.26364612

**Authors:** Kate Miles, Noelle Schlenk, Meiqian Ma, Bahare Farhadian, Samantha Leonard, Olivia Knight, Sai Alekha Challa, Shawn Gao, Samuel H. Gunther, Cindy Manko, Yuhuan Xie, Paula Tran, Melissa Silverman, Kevin W. Hoffman, Margo Thienemann, Jennifer Frankovich

## Abstract

**Objectives:** Patients with pediatric acute-onset neuropsychiatric syndrome (PANS) frequently report pain and fatigue. While some patients experience resolution of pain and fatigue following improvement in psychiatric symptoms, others develop persistent symptoms resembling fibromyalgia. Understanding these trajectories is critical for early identification and intervention in patients at risk for chronic pain or fatigue. In this study we characterize fibromyalgia symptoms in PANS and identify predictors of symptom progression following a PANS flare.

**Design:** Prospective observational cohort study.

**Setting:** Stanford University Immune Behavioral Health (IBH) Clinic in Menlo Park, California, USA.

**Participants:** 204 consecutive patients with PANS evaluated by the IBH Clinic. The average (SD) age at first neuropsychiatric deterioration was 8.2 (3.3) years.

**Main outcome measures:** We prospectively collected data on pain, fatigue, and other fibromyalgia symptoms using the 2016 American College of Rheumatology (ACR) Fibromyalgia Survey Questionnaire (FSQ), which includes the Widespread Pain Index (WPI) and Symptom Severity Scale (SSS). For the reference standard, clinician-documented diagnoses of pain syndromes were ascertained through a retrospective review of participants’ electronic medical records. We report the prevalence of these symptoms in PANS patients and controls and evaluate factors associated with time to meeting ACR fibromyalgia criteria threshold scores following clinic presentation using a Cox proportional hazards model.

**Results:** Among 204 patients with PANS (110 boys [54%]), eighty-five (42%) met ACR fibromyalgia criteria threshold scores at least once during follow-up (95% CI at 10 years follow-up time: 60.5 [46.7-74.5]). Twenty-eight of these 85 patients (13.7% of total cohort) progressed to meet full ACR fibromyalgia criteria (95% CI at 10 years follow-up time: 24.1 [15.6-36.2]). Compared with controls, patients with PANS reported substantially elevated rates of pain, daytime fatigue, waking unrefreshed, and cognitive symptoms (“brain fog”). Higher scores on the WPI and SSS at presentation were associated with a higher hazard of subsequently meeting ACR fibromyalgia criteria threshold scores.

**Conclusions:** Fibromyalgia symptoms were common in this PANS cohort, with a notable subset developing symptoms meeting full ACR fibromyalgia criteria. These findings suggest that pain and fatigue may represent post-inflammatory sequelae of PANS and may overlap mechanistically with other post-infectious conditions. Recognizing these symptom patterns may inform clinical management, development of outcome measures, and future clinical trials in these patients, as well as support investigation into shared neuroimmune mechanisms underlying post-infectious pain and fatigue syndromes.

**Key Points:** *Question:* In youth with pediatric acute-onset neuropsychiatric disorder (PANS), what is the prevalence, severity, and clinical pattern of fibromyalgia and fibromyalgia symptoms (pain, fatigue, brain fog, sleep disturbance, exercise intolerance, sensory amplification, etc.)? Which baseline clinical and demographic factors predict the subsequent fulfillment of diagnostic criteria for fibromyalgia in this population?

*Findings:* Among 204 consecutive patients with PANS evaluated at a single center, pain and fatigue were common at presentation and persisted in a subset of patients. Forty-two percent of subjects met American College of Rheumatology (ACR) fibromyalgia criteria threshold scores at least once during follow-up, with fourteen percent ultimately progressing to meet full ACR fibromyalgia criteria.

*Meaning:* Pain and fatigue may represent long-term sequelae in PANS, overlapping with other hypothesized post-infectious syndromes like myalgic encephalomyelitis/chronic fatigue syndrome (ME/CFS) and Long coronavirus disease (“Long COVID”). Acknowledging this symptomatic overlap is crucial for stimulating research into shared neuroimmune mechanisms and may reveal a common post-infectious pathophysiology.

## Introduction

Pediatric acute-onset neuropsychiatric syndrome (PANS) is characterized by the abrupt onset of obsessive-compulsive disorder (OCD) and/or eating restriction accompanied by additional neuropsychiatric symptoms. PANS typically follows a relapsing-remitting course oscillating between escalated neuropsychiatric symptoms with heightened functional impairment (“flares”) and periods of significant improvement with little to no residual symptoms (“recoveries”).^1^ Patients evaluated for PANS in the Stanford University Immune Behavioral Health (IBH) Clinic frequently report pain and fatigue; however, these symptoms are often overshadowed by severe neuropsychiatric symptoms at initial presentation. In some patients, pain and fatigue persist beyond the improvement of other neuropsychiatric symptoms. Despite these observations, little is known about the burden and characteristics of pain and fatigue in patients with PANS.

Fibromyalgia is a pain processing syndrome characterized by widespread musculoskeletal pain persisting for three or more months, accompanied by daytime fatigue, waking unrefreshed, cognitive symptoms (“brain fog”), headaches, pain/cramps in abdomen, and/or depression.^2–4^ Neuroimaging research has suggested that patients with fibromyalgia may have altered neuronal activity in brain regions implicated in pain and sensory processing, consistent with central nervous system sensitization.^5^ Altered basal ganglia activity–which is believed to play a key role in the processing of pain and sensory signals–has been implicated in both fibromyalgia and PANS and may represent shared circuitry between the two conditions.^6–14^ Emerging evidence—including work from the Karolinska Institute—indicates that inflammatory processes (marked by elevated pro-inflammatory cytokines) may contribute to the development and maintenance of fibromyalgia symptoms.^15–18^ While fibromyalgia lacks a known organic root cause, it is reported at high rates in inflammatory diseases including arthritis (psoriatic [PsA], rheumatoid [RA], and spondyloarthritis [SpA]) and systemic lupus erythematosus (SLE).^19–21^ Converging evidence suggests that PANS is also an inflammatory condition which involves pro-inflammatory monocytes, autoantibodies targeting basal ganglia interneurons, elevated cytokines, and high rates of enthesitis, vasculitis, and comorbid autoimmune diseases including arthritis.^8,22–24^ Recent advances in fibromyalgia genetics further support this conceptual framework and reinforce the concept of fibromyalgia as a central nervous system disorder with peripheral immune or neuroimmune mechanisms acting as disease triggers in susceptible individuals.^25^

Given our observations that pain and fatigue are common among patients in our PANS cohort, we sought to systematically characterize these and other fibromyalgia symptoms in this population. We describe the prevalence of fibromyalgia among these patients as per the criteria set forth by the American College of Rheumatology (ACR) and identify risk factors for developing fibromyalgia symptoms after a PANS flare using a Cox proportional hazards model.^2^

## Methods

### Study Design and Participants

We conducted a longitudinal cohort study of 204 consecutive patients with a diagnosis of PANS.^26,27^ Patients were evaluated and followed at the Stanford University IBH Clinic between 15 September 2012 and 1 January 2026 (**eFigure 1**). Patients are referred to the IBH Clinic if they experience a psychiatric or behavioral deterioration not attributable to psychosocial stressors. To maximize the likelihood of PANS diagnosis, patients evaluated by the IBH Clinic must first pass a rigorous screening protocol which prioritizes entry for local patients who have a high probability of having PANS based on medical record review with or without discussion with referring and/or primary care physician (**eAppendix 1**). Approximately 10% of referrals are accepted based on this screening process. The average (SD) age at presentation to the IBH Clinic was 10.1 (3.3) years. We also present data from 111 community control youth recruited from the same region (**eAppendix 2**).

The analysis utilized prospectively collected serial assessments using the ACR Fibromyalgia Survey Questionnaire (FSQ) (**eAppendix 3**) and additional symptom data (sleep disturbance, exercise intolerance, sensory amplification), demographic data, and medical record data (clinical documentation of pain syndromes and comorbid immune disorders).^2^

The Stanford Institutional Review Board (IRB) approved this prospective study protocol (IRB#26922), which included consent to review all electronic medical record data. Adults and parents of minors gave informed written consent, and competent patients aged 7-17 gave assent. Study data were recorded and managed in REDCap (**eAppendix 4**). Study data were extracted for analysis on 14 June 2026 and censored as of 31 December 2025. Sex and race/ethnicity were self-reported and included to assess generalizability. Race/ethnicity categories used were those from the Office of Management and Budget (2024 revision). Geographic regions were categorized according to the California Census 2020 regional definitions.

### Patient and Public Involvement

Patients and/or caregivers are asked to fill out an IBH questionnaire before each clinical encounter. These questionnaires were originally designed in 2012 and evolved over time in response to patient, caregiver, and clinician concerns about specific emerging symptoms and comorbidities. Questionnaire components based on patient and caregiver feedback—including the metrics of pain and malaise reported in this study—are helpful in guiding ongoing PANS research.

### Assessment of Pain, Daytime Fatigue, Waking Unrefreshed, Cognitive Symptoms (Brain Fog), and Other Fibromyalgia Symptoms

The FSQ gathers data on pain location (checklist of 19 specific body areas), pain regions, severity of additional symptoms (daytime fatigue, waking unrefreshed, cognitive symptoms [“brain fog”]) and presence of other somatic symptoms (headaches, abdominal pain, and depression) (**eAppendix 3**). Patients meet ACR fibromyalgia criteria threshold scores when their combination of FSQ scores (Widespread Pain Index [WPI] and Symptom Severity Scale [SSS]) fulfill the third criterion of the 2016 ACR fibromyalgia criteria (**eAppendix 3**). Patients meet full ACR fibromyalgia criteria when the questionnaire respondent(s) additionally report generalized pain (first criterion), that has persisted at a similar level for three or more months (second criterion). The presence of generalized pain—defined as pain in at least 4 of 5 regions of the body (third criterion)—is determined from the WPI (**eAppendix 3**). For each patient meeting the first and third criterion, we conducted chart review of the surrounding 3-month period to establish whether the patient’s fibromyalgia symptoms were present at a similar level for 3 or more months (thus fulfilling the second criterion). If FSQ scores were unavailable during this 3-month period of clinical history, we reviewed the electronic medical record (EMR) for other documentation of symptom persistence. In the absence of scores or clear documentation of symptoms, we did not consider the patient to have fulfilled the second criterion.

The first IBH Clinic visit at which all three ACR fibromyalgia criteria were fulfilled is defined as the time point meeting “full ACR fibromyalgia criteria.” We recognize that some patients may experience fibromyalgia symptoms before presentation to the IBH Clinic.

However, as patients do not consistently complete the FSQ prior to their first visit, we cannot confidently determine their satisfaction of the ACR fibromyalgia criteria prior to their first visit.

Sleep disturbance was assessed using the PANS 31-Item Scale.^28^ Because earlier versions of this instrument recorded sleep disturbance as a dichotomous (yes/no) item, whereas later versions used severity ratings, sleep disturbance was considered present if the item was endorsed (“yes”) or assigned a score greater than 0. Additional fibromyalgia symptom variables (including exercise intolerance and sensory disturbances) were derived from harmonized questionnaire items. Some items were added over time, and minor differences in wording and response format were accounted for during harmonization. The availability of these variables across participants is described in the table footnotes.

### Clinical Documentation of Pain Syndromes

A research specialist (KM) reviewed the EMR—including clinical notes, problem lists, and other physician documentation—for clinically documented pain diagnoses using the following search terms: amplified musculoskeletal pain, amplified abdominal or head pain, functional abdominal pain, amplified pain or primary pain disorder, central pain sensitivity/dysregulation, chronic pain syndrome, complex regional pain syndrome (CRPS), diffuse myofascial pain, widespread pain, electrical shock-like pain, and fibromyalgia.

### Classifications of Comorbid Conditions

Pediatric rheumatologists (MM, JF) classified patients with arthritis if they met ACR criteria for juvenile idiopathic arthritis (JIA) (**eAppendix 5**). Diagnoses of other autoimmune conditions including celiac disease, psoriasis, thyroiditis, inflammatory bowel disease (IBD) (including common variable immunodeficiency [CVID] associated IBD) and SLE were assigned based on sub-specialist evaluation (**eAppendix 5**). Separately, we evaluated other immunologic conditions including eosinophilic esophagitis (EoE), primary immunodeficiency (PID), and chronic urticaria (**eAppendix 5**).

### Statistical Analyses

We estimated the cumulative incidence of patients meeting ACR fibromyalgia criteria threshold scores and calculated 95% CIs using product-limit (Kaplan-Meier) survival analysis methods to account for censoring.

We applied a Cox proportional hazards regression model to examine the risk of meeting ACR fibromyalgia criteria threshold scores. Covariates were selected *a priori* based on clinical relevance and include age at first IBH Clinic visit, sex, disease course at clinic presentation, presence of sleep disturbance(s) at intake (based on first completed 31-Item PANS Scale within 90 days of presentation), WPI at intake (within 90 days of presentation), and SSS at intake (within 90 days of presentation). Patients without available scores at intake were excluded from this model (n=19 excluded for absence of full FSQ [WPI and/or SSS]; n=17 excluded for absence of 31-Item PANS Scale). We used standard procedures to check model assumptions including multicollinearity.

We analyzed data using Python 3.10 (*lifelines, pandas*, *NumPy*, *SciPy*).^29–31^ We used *matplotlib* and *seaborn* implemented in Python 3.10 to generate plots.^32,33^ The analysis code is available in a public GitHub repository (n-schlenk-som/pain-fatigue-pans).

## Results

The analyses included data from 204 patients who met PANS criteria 110 boys [53.9%]; 151 White or European [74.0%]) and 111 controls (57 boys [51.4%]; 54 White or European [48.6%]) (**Table 1**). We report descriptive statistics for clinical features at intake (**eTable 1**).^28^ In the PANS cohort, the mean (SD) age at PANS onset was 9.5 (3.4) years and the mean (SD) follow-up time was 5.2 (3.2) years (time from initial clinical encounter to most recent encounter as of 31 December 2025). The mean (SD) age of controls at first clinic visit with available data was 12.0 (3.5) years. We do not follow controls longitudinally.

**Table 1.** Demographics and Neuropsychiatric Presentation of Consecutive Patients with PANS (N=204) and Controls (N=111) Seen at a Single Center.

| Characteristic | Patients (N=204) | Controls (N=111) |
| --- | --- | --- |
| Sex at birth, No. (%) |  |  |
| Female | 94 (46.1) | 57 (51.4) |
| Male | 110 (53.9) | 54 (48.6) |
| Race, No. (%) |  |  |
| White or European | 151 (74.0) | 54 (48.6) |
| Asian or Asian American | 6 (2.9) | 32 (28.8) |
| Hispanic or Latino (may include White) | 17 (8.3) | 5 (4.5) |
| Multiracial or Other | 30 (14.7) | 20 (18.0) |
| Age at first-ever neuropsychiatric deterioration, mean (SD), years | 8.2 (3.3) | - |
| Age at deterioration prompting referral to clinic, mean (SD), years | 9.6 (3.2) | - |
| Age at PANS onset, mean (SD), years | 9.5 (3.4) | - |
| Age at IBH Clinic presentation, mean (SD), years | 10.1 (3.3) | 12.0 (3.5) |
| Meets PANS criteria at or before IBH Clinic presentation, No. (%) | 178 (87.3) | - |
| Clinical course<sup>a</sup> at IBH Clinic presentation, No. (%) |  |  |
| Never persistent | 184 (90.2) | - |
| Ever persistent | 20 (9.8) | - |
| IBH Clinic follow-up time, mean (SD), years | 5.2 (3.2) | - |
| Clinical course<sup>a</sup> at time of study, No. (%) |  |  |
| Single flare<sup>b</sup> | 21 (10.3) | - |
| Relapsing-remitting<sup>c</sup> | 178 (87.3) | - |
| Primary persistent | 0 | - |
| Secondary persistent | 5 (2.5) | - |

<sup>a</sup>We classified patients into their respective clinical courses at clinical presentation and at the time of study using definitions proposed by Masterson et al. (2025). *Single flare*: One recorded psychiatric flare to date. If flare is ongoing (i.e., without an unequivocal recovery), duration of flare must be <12 months at time of study. *Relapsing-remitting*: At least one unequivocal recovery and subsequent psychiatric flare start. If flare is ongoing, duration of flare must be <12 months at time of study. *Primary persistent*: One recorded psychiatric flare to date with duration of flare ≥12 months without an unequivocal recovery and ongoing at time of study. *Secondary persistent*: A prior relapsing-remitting course with subsequent psychiatric flare ≥12 months without an unequivocal recovery and ongoing at time of study. Psychiatric flare information was extracted retrospectively from the EMR.
<sup>b</sup>4/17 patients classified as *single flare* experienced a persistent flare.
<sup>c</sup>60/178 patients classified as *relapsing-remitting* have experienced at least one persistent flare to date.
**Abbreviations:** Pediatric acute-onset neuropsychiatric syndrome (PANS); Immune Behavioral Health (IBH); electronic medical record (EMR).

Most patients reported pain not due to injury, daytime fatigue, waking unrefreshed, cognitive symptoms (“brain fog”), sleep disturbance, exercise intolerance, and sensory disturbances at least once during the first year of follow-up (**eTable 2**). At the visit closest to the one-year follow-up date, patients continued to report pain not due to injury, daytime fatigue, waking unrefreshed, cognitive symptoms (“brain fog”), headaches, pain/cramps in abdomen, depression, sleep disturbance, exercise intolerance, and sensory disturbances (**Table 2, eTable 3**). These symptoms were common during PANS flares and, in some patients, persisted beyond partial and full neuropsychiatric recovery (**Figure 1**).

**Figure 1.**
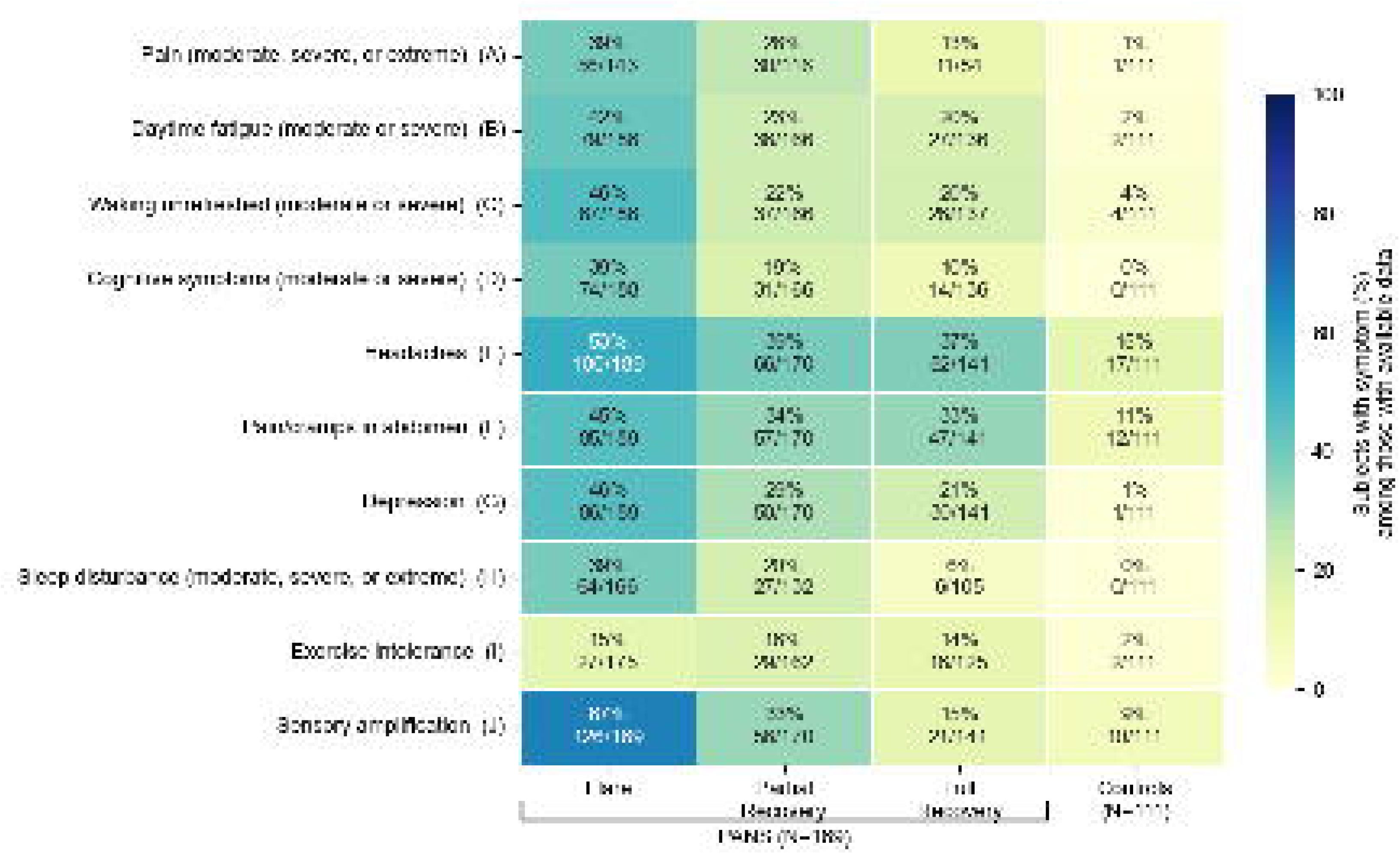
Prevalence of Pain, Fatigue, Cognitive, Sleep, Sensory and Other Symptoms During Flare and Recovery Across Consecutive Patients with PANS (N=186) and Controls (N=111) Prevalence of fibromyalgia symptoms across 189 patients with PANS in different clinical states (PANS flare, partial recovery, full recovery) and 111 controls. Items (A) and (H) were derived from the 31-Item PANS Scale, which measures symptoms using a 5-point Likert-type response scale ranging from 0 (none) to 4 (extreme). Items (B) through (D) were derived from the section of the FSQ which measures symptom severity with a 4-point Likert-type response scale ranging from 0 (none) to 3 (severe). Items (E) through (G) were derived from the section of the FSQ which captures symptoms as either present or absent. Items (I) and (J) are part of the IBH Clinic questionnaire and are captured as either present or absent. Fifteen patients from our study cohort (N=204) are excluded because they did not have any questionnaires from a PANS flare. For each patient and symptom, the *Flare* column represents data from the patient’s first PANS flare for which that symptom was assessed. The *Partial Recovery* and *Full Recovery* columns represent data from the first partial recovery and full recovery visits after the flare, respectively, at which that symptom was assessed. Percentages are calculated from the number of patients with available data for the respective symptom and clinical state. Control data are taken from the first visit at which the WPI, SSS, and ancillary symptoms were collected. *Abbreviations*: Pediatric acute-onset neuropsychiatric syndrome (PANS); Fibromyalgia Survey Questionnaire (FSQ); Widespread Pain Index (WPI); Symptom Severity Scale (SSS).

**Table 2.**
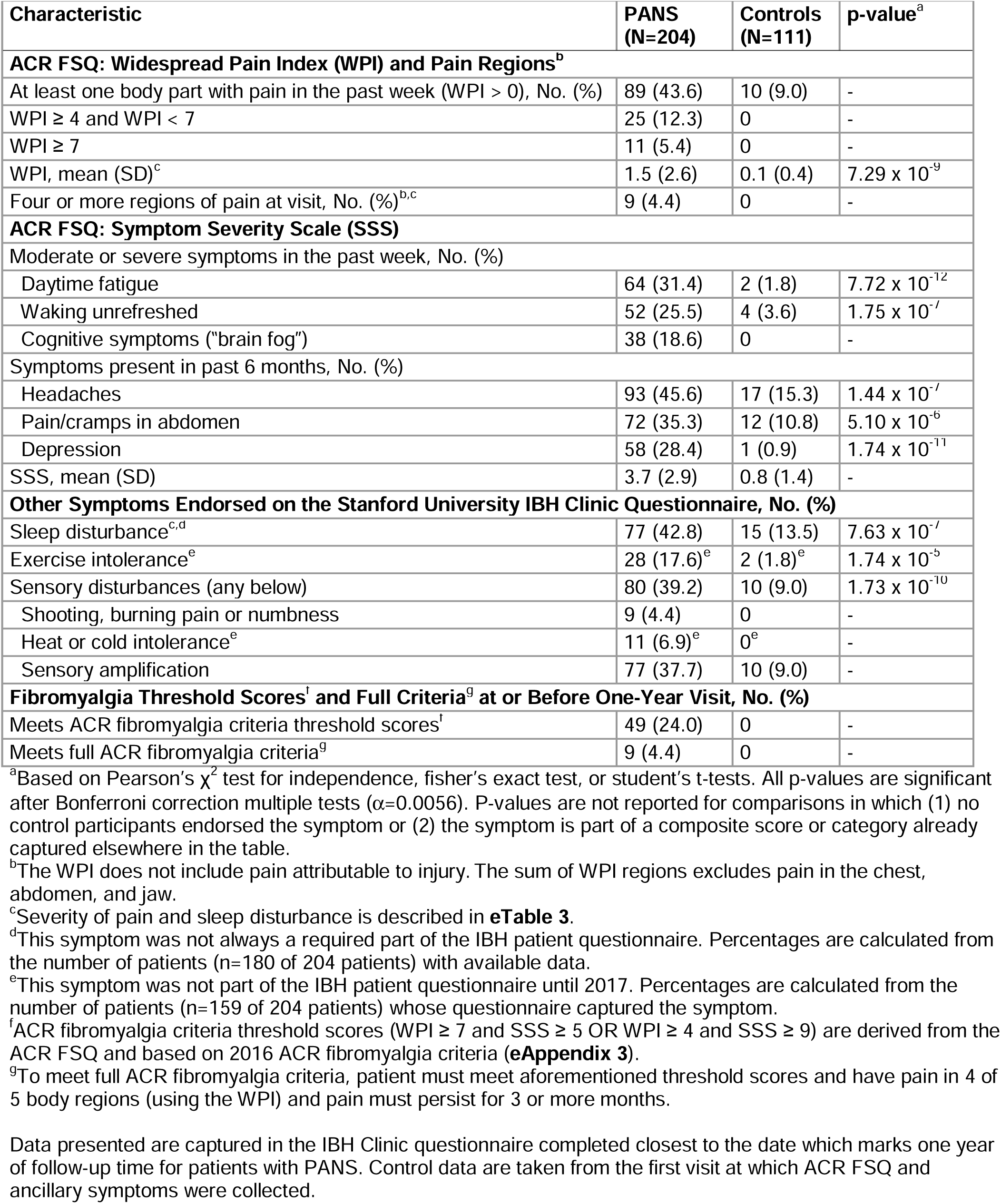
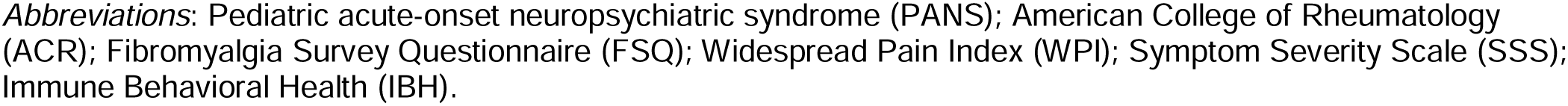
Characteristics of Pain, Fatigue, Cognitive, Sleep, and Sensory Symptoms Reported at One-Year Follow-Up Visit in Consecutive Patients with PANS (N=204) and at First Visit in Controls (N=111)

In the first year of follow-up, 46 of 199 patients with available first-year data met ACR fibromyalgia criteria threshold scores (23.1%), corresponding to a Kaplan-Meier estimated cumulative incidence of 22.6% (**Figure 2**). Of these, 9 (4.5% [of 199 with available data]) progressed to meet full ACR fibromyalgia criteria within their first year of follow-up.

**Figure 2.**
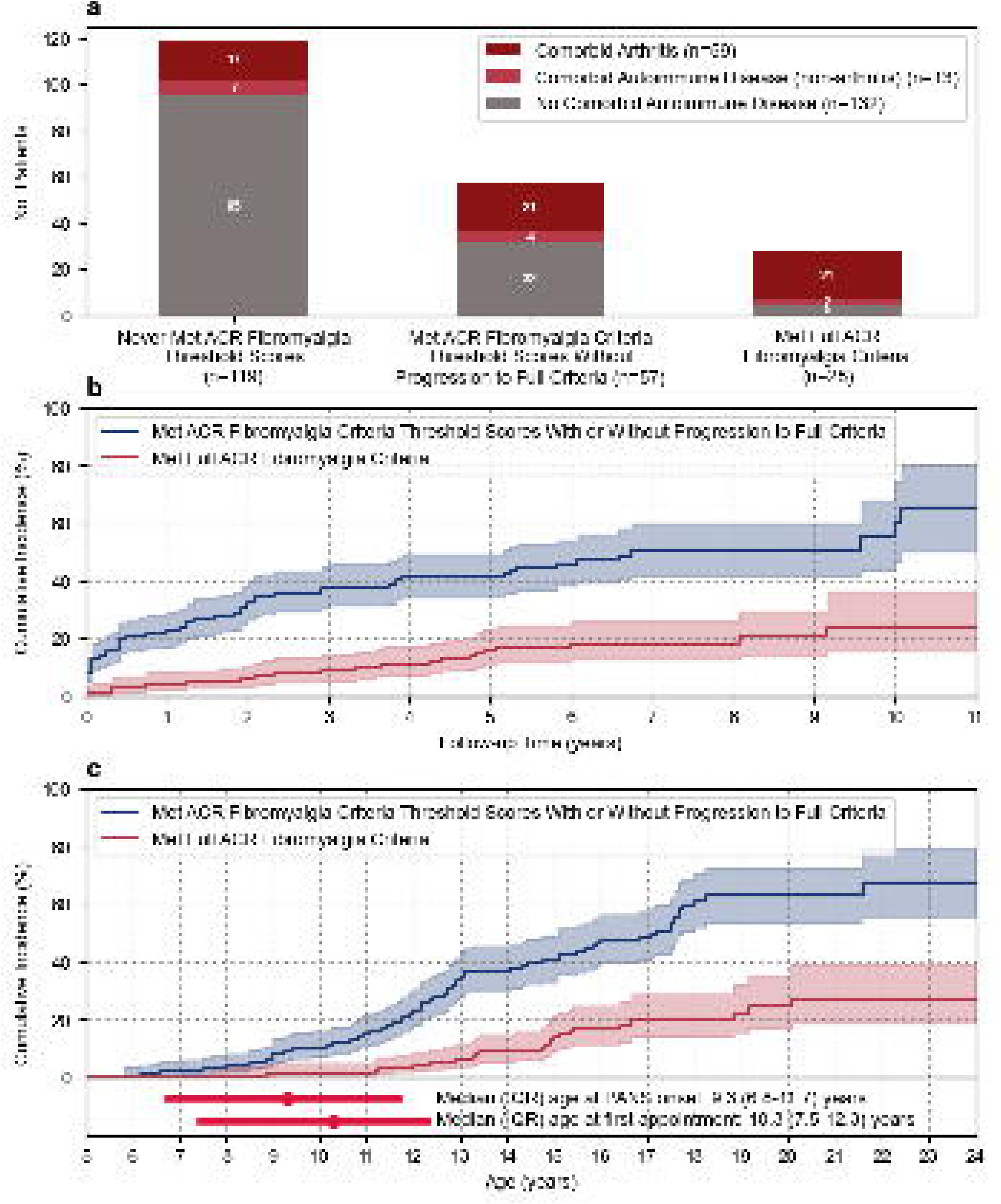
Frequency and Cumulative Incidence of Meeting ACR Fibromyalgia Criteria Threshold Scores and Full ACR Fibromyalgia Criteria Among Consecutive Patients with PANS (N=204) **(a)** Frequencies of comorbid arthritis and other autoimmune diseases among 204 patients with PANS, stratified by satisfaction of ACR fibromyalgia criteria threshold scores and full criteria. **(b)** Kaplan-Meier plot of time (in years) from first IBH appointment to the appointment at which ACR fibromyalgia criteria threshold scores (blue) or full criteria (red) are met. **(c)** Kaplan-Meier plot of age (in years) at which ACR fibromyalgia criteria threshold scores (blue) or full criteria (red) are met. The shaded areas indicate the 95% CIs. Subjects are censored at their most recent IBH appointment date as of 31 December 2025. ACR fibromyalgia criteria threshold scores (WPI ≥ 7 and SSS ≥ 5 OR WPI ≥ 4 and SSS ≥ 9) are derived from the ACR FSQ and based on 2016 ACR fibromyalgia criteria (**eAppendix 3**). To meet full ACR fibromyalgia criteria, patient must meet aforementioned threshold scores and have pain in 4 of 5 body regions (using the WPI) and pain must persist for 3 or more months. *Abbreviations*: American College of Rheumatology (ACR); Pediatric acute-onset neuropsychiatric syndrome (PANS); Confidence interval (CI); interquartile range (IQR); Widespread Pain Index (WPI); Symptom Severity Scale (SSS); Fibromyalgia Survey Questionnaire (FSQ).

Eighty-five patients (41.7%) met ACR fibromyalgia criteria threshold scores (WPI and SSS) at least once during their follow-up by the IBH Clinic (**Figure 2**, **Table 3**). Of these, 28 (32.9%; 13.7% of total cohort) progressed to meet full ACR fibromyalgia criteria at some point during their follow-up period.

**Table 3.** Prevalence of Fibromyalgia in Consecutive Patients with PANS (N=204), Evaluated Per American College of Rheumatology (ACR) Fibromyalgia Criteria.

| Characteristic | Patients (N=204) |
| --- | --- |
| **Ever met ACR fibromyalgia criteria threshold scores (WPI and SSS), No. (%)** | 85 (41.7) |
| **Met ACR fibromyalgia criteria threshold scores without progression to full ACR fibromyalgia criteria, No. (%)** | **57 (27.9)** |
| Age at meeting ACR fibromyalgia criteria threshold scores, mean (SD), years | 12.4 (3.7) |
| Time from clinic presentation to meeting ACR fibromyalgia threshold scores, mean (SD), years | 1.9 (2.6) |
| **Met full ACR fibromyalgia criteria, No. (%)** | **28 (13.7)** |
| Age at meeting ACR fibromyalgia criteria threshold scores, mean (SD), years | 12.3 (2.4) |
| Time from clinic presentation to meeting ACR fibromyalgia threshold scores, mean (SD), years | 1.1 (1.5) |
| Age at meeting full ACR fibromyalgia criteria, mean (SD), years | 13.9 (3.0) |
| Time from clinic presentation to full ACR fibromyalgia criteria, mean (SD), years | 2.8 (2.5) |

ACR fibromyalgia criteria are based on assessments of pain (WPI) and other symptoms like waking unrefreshed, daytime fatigue, and cognitive symptoms ("brain fog") (SSS) (**eAppendix 3**). ACR fibromyalgia criteria threshold scores (WPI $\geq 7$ and SSS $\geq 5$ OR WPI $\geq 4$ and SSS $\geq 9$ ) are derived from the ACR FSQ and based on 2016 ACR fibromyalgia criteria. To meet full ACR fibromyalgia criteria, patient must meet aforementioned threshold scores and have pain in 4 of 5 body regions (using the WPI) and pain must persist for 3 or more months.
*Abbreviations:* Pediatric acute-onset neuropsychiatric syndrome (PANS); American College of Rheumatology (ACR); Widespread Pain Index (WPI); Symptom Severity Scale (SSS); Fibromyalgia Survey Questionnaire (FSQ).

We report fibromyalgia symptoms at the time of meeting ACR fibromyalgia criteria threshold scores for 28 patients meeting full ACR fibromyalgia criteria and 57 who met ACR fibromyalgia criteria threshold scores (WPI and SSS) but did not progress to meet full ACR fibromyalgia criteria (pain was not generalized or did not persist three or more months) (**eTable 4**). At this timepoint, 56 patients (65.9% [56 of 85]) report some type of sensory disturbance.

Patients most frequently reported pain in the axial region, which includes the neck (57.6% [49 of 85]) and back (72.9% [62 of 85]). Other common findings include leg (76.5% [65 of 85]) and shoulder (49.4% [42 of 85]) pain. Forty-one patients (48.2% [41 of 85]) had at least four regions of pain involved. The average (SD) number of pain sites (out of 19) was 7.8 (3.2).

We report clinical characteristics, ACR FSQ and other psychometric scores at presentation, and comorbid immune disorders (arthritis, other autoimmune disease, and other immunologic conditions), stratified by satisfaction of ACR fibromyalgia criteria threshold scores and progression to meeting full ACR fibromyalgia criteria (**eTable 5**). We report time to meeting ACR fibromyalgia criteria threshold scores and full criteria as functions of follow-up time and age (**Figure 2**, **eTables 6-7**).

Documentation of a pain syndrome was common among patients meeting ACR fibromyalgia criteria threshold scores, especially those who progressed to meet full ACR fibromyalgia criteria. Among 57 patients meeting ACR fibromyalgia criteria threshold scores who did not progress to meet full ACR fibromyalgia criteria, 25 (43.9%) had at least one clinically documented pain syndrome. Most patients meeting full ACR fibromyalgia criteria had a documented pain syndrome (82.1% [23 of 28]). Eight patients who did not meet ACR fibromyalgia criteria threshold scores had a documented pain syndrome (6.7% [8 of 119]). We report frequencies of patients with each clinically documented pain syndrome term (**Table 4**) and discuss reasons for discrepancies between clinical diagnoses and ACR fibromyalgia criteria results (**eAppendix 6**). We show overlapping longitudinal trajectories of ACR fibromyalgia criteria threshold scores, satisfaction of ACR fibromyalgia criteria threshold scores, satisfaction of full ACR fibromyalgia criteria, and clinical documentation of pain syndromes in a swimmer plot (**Figure 3**). We similarly show per-subject longitudinal trajectories of the WPI and each major component of the SSS (waking unrefreshed, daytime fatigue, and cognitive symptoms [“brain fog”]) (**eFigures 3-6**).

**Figure 3.**
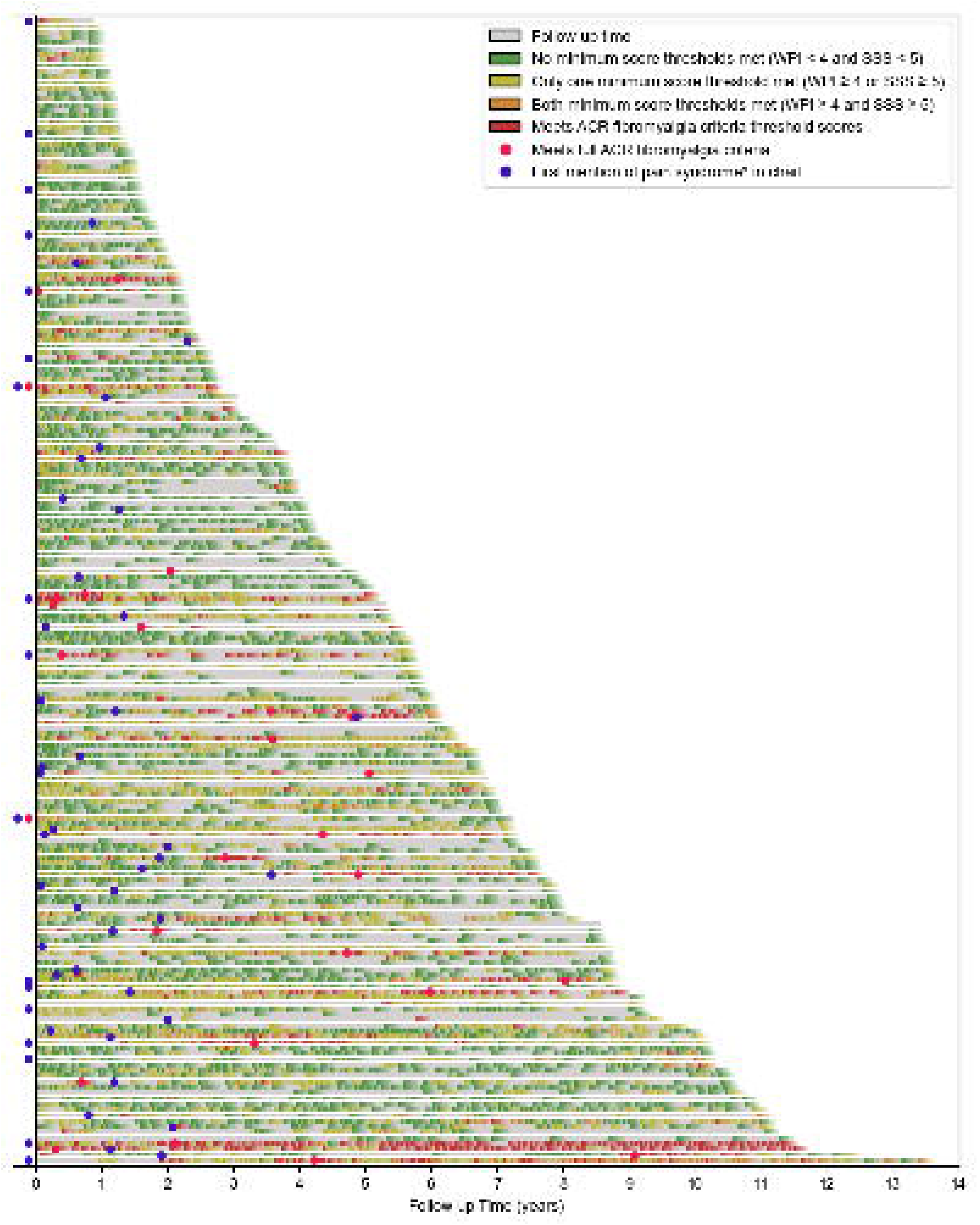
Longitudinal Trajectories of Fibromyalgia Symptom Scores and Diagnoses in Consecutive Patients with PANS (N=204) Swimmer plot depicting 204 patient trajectories over the course of follow-up by the IBH Clinic. Colored segments (green, yellow, orange) denote satisfaction of minimum threshold scores collected at patient visits. Red segments indicate a visit at which the subject meets ACR fibromyalgia criteria score thresholds. Per-visit events are marked by the start of a color, followed by a fading line with length 90 days unless interrupted by a subsequent visit. Patients are ordered by duration of follow-up. ACR fibromyalgia criteria threshold scores (WPI ≥ 7 and SSS ≥ 5 OR WPI ≥ 4 and SSS ≥ 9) are derived from the ACR FSQ and based on 2016 ACR fibromyalgia criteria (**eAppendix 3**). To meet full ACR fibromyalgia criteria, patient must meet aforementioned threshold scores and have pain in 4 of 5 body regions (using the WPI) and pain must persist for 3 or more months. * *Pain syndrome* reflects a documentation in the electronic medical record the diagnoses or mention of any term reflecting pain amplification including 1) regional pain syndrome (e.g. functional abdominal pain), 2) evidence of amplified pain on physical exam (e.g., myofascial point tenderness or pain which is out of proportion to physical findings of arthritis or injury) and 3) widespread pain (e.g., fibromyalgia). Situations 1 and 2 may prompt clinician suspicion of amplified pain even in cases where the patient/caregiver does not report threshold scores using the ACR FSQ. Thus, clinician documentation of pain syndrome does not always overlap with meeting ACR fibromyalgia criteria (see **eAppendix 3** for case details). See **eFigures 3-6** for separation of fibromyalgia components (pain, daytime fatigue, waking unrefreshed, cognitive symptoms [“brain fog”]). *Abbreviations*: Pediatric acute-onset neuropsychiatric syndrome (PANS); Immune Behavioral Health (IBH); American College of Rheumatology (ACR); Widespread Pain Index (WPI); Symptom Severity Scale (SSS); Fibromyalgia Survey Questionnaire (FSQ).

**Table 4.** Descriptive Terms Reflecting Pain Syndrome Documented in Clinical Notes for Consecutive Patients with Patients with PANS, Stratified by Satisfaction of American College of Rheumatology (ACR) Fibromyalgia Criteria (N=204)

|  | Never Met ACR Fibromyalgia Criteria Threshold Scores (WPI and SSS)<sup>a</sup> | Met ACR Fibromyalgia Criteria Threshold Scores (WPI and SSS)<sup>a</sup><br>*No progression to full ACR fibromyalgia* | Met Full ACR Fibromyalgia Criteria<sup>a</sup> |
| --- | --- | --- | --- |
| **No. Patients** | **119** | **57** | **28** |
| **Any mention of a pain syndrome or diagnosis in chart, No. (%)** | 8 (6.7)<sup>b</sup> | 25 (43.9) | 23 (82.1) |
| Amplified musculoskeletal pain (AMPS) | 0 | 0 | 3 (10.7) |
| Amplified pain involving the abdomen or functional abdominal pain | 1 (0.8) | 3 (5.3) | 3 (10.7) |
| Amplified pain (other) or primary pain disorder | 3 (2.5) | 13 (22.8) | 17 (60.7) |
| Central (pain) sensitivity pain processing disorder, pain dysregulation, pain syndrome | 5 (4.2) | 12 (21.1) | 11 (39.3) |
| Diffuse myofascial pain | 0 | 0 | 1 (3.6) |
| Diffuse, widespread, generalized, or whole-body pain (syndrome) | 4 (3.4) | 10 (17.5) | 19 (67.9) |
| Complex regional pain syndrome (CRPS)<sup>c</sup> | 0 | 1 (1.8) | 1 (3.6) |
| Electrical shock-like pain<sup>c</sup> | 0 | 7 (12.3) | 13 (46.4) |
| Fibromyalgia<sup>d</sup> | 0 | 5 (8.8) | 13 (46.4) |

<sup>a</sup> ACR fibromyalgia criteria threshold scores (WPI $\geq 7$ and SSS $\geq 5$ OR WPI $\geq 4$ and SSS $\geq 9$ ) are derived from the ACR FSQ and based on 2016 ACR fibromyalgia criteria (**eAppendix 3**). To meet full ACR fibromyalgia criteria, patient must meet aforementioned threshold scores and have pain in 4 of 5 body regions (using the WPI) and pain must persist for 3 or more months.
<sup>b</sup> We describe these cases in **eAppendix 6**.
<sup>c</sup> This term was only recorded if it was used in the context of describing the entire pain syndrome (not one-off instances).
<sup>d</sup> Although clinicians had the results of the ACR FSQ, they may not have documented the diagnosis of fibromyalgia in the clinic notes (especially in very young children) but rather used a different pain term if they felt it was more appropriate for the case and/or discussions with the family.
**Abbreviations:** American College of Rheumatology (ACR); Widespread Pain Index (WPI); Symptom Severity Scale (SSS); Fibromyalgia Survey Questionnaire (FSQ).

In the exploratory Cox proportional hazards model, two scores at baseline were independently associated with a higher hazard of meeting ACR fibromyalgia criteria threshold scores: WPI (adjusted hazard ratio [aHR], 1.40 95% CI, 1.26-1.55; p<0.005) and SSS (aHR, 1.38; 95% CI, 1.24-1.55; p<0.005). Sex, age at clinical presentation, history of persistent disease course at clinic presentation, and presence of sleep disturbance at clinic presentation were not significantly associated with time to meeting ACR fibromyalgia criteria threshold scores (**Table 5**, **eTables 8**-**9**).

**Table 5.** Exploratory Cox Proportional Hazards Model of Time to Meeting American College of Rheumatology (ACR) Fibromyalgia Criteria Threshold Scores Among Consecutive Patients with PANS (N=168)

|  | Never Met ACR<br>Fibromyalgia Criteria<br>Threshold Scores (WPI<br>and SSS)<sup>a</sup> | Met ACR Fibromyalgia Criteria<br>Threshold Scores (WPI and SSS)<sup>a</sup><br>*With or without progression to full*<br>*ACR fibromyalgia* | Cox Proportional Hazards<br>Model<sup>b</sup> |  |
| --- | --- | --- | --- | --- |
| No. | 104 | 64 |  |  |
| Covariates | No. (%) or median [IQR] | No. (%) or median [IQR] | aHR (95% CI) | p-value |
| Sex (male) | 54 (51.9) | 36 (56.2) | 1.10 (0.64-1.90) | 0.73 |
| Age at clinical presentation, years | 9.4 [6.4, 12.0] | 10.9 [9.0, 12.6] | 1.04 (0.95-1.14) | 0.37 |
| Ever persistent at clinic intake | 5 (4.8) | 6 (9.4) | 0.87 (0.35-2.13) | 0.76 |
| WPI at intake | 0.0 [0.0, 2.0] | 2.0 [1.0, 5.2] | 1.40 (1.26-1.55) | <0.005 |
| SSS at intake | 4.0 [2.0, 6.0] | 7.0 [5.0, 9.0] | 1.38 (1.24-1.55) | <0.005 |
| Sleep disturbance at intake | 50 (48.1) | 40 (62.5) | 1.53 (0.88-2.65) | 0.13 |

<sup>a</sup>ACR fibromyalgia criteria threshold scores (WPI $\geq 7$ and SSS $\geq 5$ OR WPI $\geq 4$ and SSS $\geq 9$ ) are derived from the ACR FSQ and based on 2016 ACR fibromyalgia criteria (**eAppendix 3**). To meet full ACR fibromyalgia criteria, patient must meet aforementioned threshold scores and have pain in 4 of 5 body regions (using the WPI) and pain must persist for 3 or more months.
<sup>b</sup>Multivariable model. For per-covariate univariable analyses, refer to **eTable 8**.
**Abbreviations:** American College of Rheumatology (ACR); Widespread Pain Index (WPI); Symptom Severity Scale (SSS); interquartile range (IQR); adjusted hazard ratio (aHR); Fibromyalgia Survey Questionnaire (FSQ).

## Discussion

In this study of 204 patients with PANS, we observed that fibromyalgia symptoms (pain, fatigue, waking unrefreshed, cognitive symptoms [“brain fog”], sleep disturbance, exercise intolerance, and sensory disturbances) are common during acute neuropsychiatric deteriorations and persist in a subset of patients (**Figure 1**).

At the one-year follow-up visit, patients with PANS reported markedly higher rates of pain, fatigue, and other fibromyalgia symptoms compared with controls (**Table 2**). The prevalence of these symptoms greatly exceeds general pediatric population estimates, where chronic or recurrent pain is reported to affect roughly 20-25% of children and severe fatigue is reported in fewer than 5%.^34,35^ Eighty-five patients (41.7%) met the ACR fibromyalgia criteria threshold scores (WPI and SSS) at least once during follow-up, and 28 patients (13.7% of the cohort) progressed to meet full ACR fibromyalgia criteria, indicating that while many patients experience partial or intermittent symptoms, a meaningful subset develops pain, fatigue, and sensory disturbances consistent with fibromyalgia. Our findings revealed that higher WPI and SSS scores at presentation were associated with higher risk of meeting ACR fibromyalgia criteria threshold scores (with or without progression to meeting full ACR fibromyalgia criteria). Together, these findings highlight the considerable burden of fibromyalgia symptoms in PANS.

### Distribution of Chronic Pain

At the time point meeting ACR fibromyalgia criteria threshold scores, PANS patients most often reported axial pain involving the neck and back, consistent with prior pediatric and adult fibromyalgia studies.^36,37^ In contrast, shoulder pain, which is prominent in adult fibromyalgia cohorts, was less pronounced in our population. The body pain reported by PANS patients tends to be symmetrically distributed (not shown).

### Overlap Between Fibromyalgia and Arthritis

Arthritis and fibromyalgia frequently co-occur across rheumatologic diseases including rheumatoid arthritis, SpA, PsA, and SLE.^19–21^ The added presence of fibromyalgia can complicate diagnosis and management of these conditions.^19^ Similarly, we have demonstrated elevated rates of ERA and SpA-spectrum disease in patients with PANS.^24^ Fibromyalgia complicates the diagnosis and management of arthritis in PANS cases, particularly when patients continue to endorse pain despite improvement in joint inflammation documented by physical exam and imaging. Further understanding of the nuances of fibromyalgia may help providers manage PANS and PANS-related arthritis more effectively.

While arthritis may contribute to pain burden in some patients, arthritis alone does not explain the high frequency of other fibromyalgia symptoms observed in this cohort. The ACR fibromyalgia criteria include symptoms beyond pain including daytime fatigue, waking unrefreshed, cognitive symptoms (“brain fog”), headaches, pain/cramps in abdomen, and depression. Beyond the ACR FSQ and fibromyalgia criteria, additional symptoms are frequent and debilitating (sleep disturbance, exercise intolerance, and sensory disturbances).^38,39^ This distinction is clinically important because pain, fatigue, and other fibromyalgia symptoms may persist in these patients even when inflammatory joint findings improve. Conversely, because arthritis and fibromyalgia commonly coexist, the presence of fibromyalgia symptoms should not delay or preclude further rheumatologic assessment among patients with persistent joint pain.

### Sex Distribution

Unlike typical fibromyalgia populations, which have strong female predominance in both pediatric and adult studies, our PANS cohort demonstrated a near-equal distribution of male and female patients. Among patients meeting ACR fibromyalgia criteria threshold scores with or without progression to full ACR fibromyalgia criteria, 56% were male. In our Cox proportional hazards model, sex was not associated with time meeting ACR fibromyalgia criteria threshold scores. These findings suggest that fibromyalgia symptom burden in PANS may occur across sexes and may not follow the demographic pattern observed in other fibromyalgia populations.^40^

### Overlap Between PANS and Fibromyalgia

The shared clinical features of PANS and fibromyalgia may reflect convergent pathophysiology involving cortico-striato-thalamo-cortical (CSTC) circuit function. Prevailing theory suggests that inflammatory or autoimmune insults disrupt the basal ganglia’s capacity to filter sensory input and select appropriate cognitive-motor programs. This dysfunction may contribute to aberrant, perseverative signaling to the thalamus, a structure implicated in PANS and fibromyalgia.^11,41,42^ The subsequent relay of these signals to the cortex establishes a maladaptive feedback loop, manifesting clinically as obsessive-compulsive symptoms, central pain sensitization, and profound central fatigue. This model also accounts for unrefreshing sleep via failed thalamic sensory gating. However, given that neuroimaging implicates regions beyond this core circuit, it is highly probable that sleep-wake disturbances are compounded by dysfunction in distinct regulatory networks, including hypothalamic centers governing circadian timing and brainstem nuclei that promote and stabilize sleep and wakefulness.

### Overlap with Myalgic Encephalomyelitis/Chronic Fatigue Syndrome (ME/CFS)

Myalgic encephalomyelitis/chronic fatigue syndrome (ME/CFS) frequently co-occurs with fibromyalgia, likely in part due to the overlap in criteria (specifically SSS) between the two conditions.^43,44^ IOM 2015 ME/CFS criteria consists of 1) daytime fatigue lasting longer than 6 months, 2) post-exertional malaise, 3) waking unrefreshed, and 4) either cognitive impairment or orthostatic intolerance. The first, third, and part of the fourth criteria are part of the ACR FSQ (SSS component). While the data for post-exertional malaise (second IOM criterion) and orthostatic intolerance (part of the fourth IOM criterion) have not always been captured by the IBH questionnaire, the elevated prevalence of non-restorative sleep (waking unrefreshed), daytime fatigue, cognitive symptoms (“brain fog”) in the patient population meeting ACR fibromyalgia criteria indicates a potential for high overlap between these two syndromes, and an intriguing area for future study.

### Impact of Immune Modulation

Patients in the IBH Clinic are frequently treated with anti-inflammatory and/or immunomodulatory therapies alongside standard of care for their psychiatric symptoms. These therapies include nonsteroidal anti-inflammatory drugs (NSAIDs), corticosteroid bursts or corticosteroid burst with taper, intravenous immunoglobulin (IVIG) and standard of care disease-modifying antirheumatic drugs (DMARDS) for arthritis. Since these therapies are often given soon after clinical presentation, they may influence progression from transient pain symptoms to full ACR fibromyalgia.^45–48^ We suspect that a fraction of patients with pain and fatigue did not progress to meet full ACR fibromyalgia criteria because of earlier treatment of underlying inflammation. However, the influence of clinical interventions is not yet clear, as a univariate logistic regression model revealed that among patients evaluated within 2 years of the neuropsychiatric deterioration prompting referral to clinic, time to clinical presentation was not a strong predictor of symptoms meeting ACR fibromyalgia criteria threshold scores (not shown).

### Low-level Systemic Inflammation and Question of Role for Infection

New research on fibromyalgia implicates inflammatory and neuroimmune mechanisms in a subset of patients.^49–51^ This hypothesis is particularly relevant in PANS, which is suspected to be driven by low-level systemic inflammation, immune dysregulation, and neuroinflammation. While the etiology of fibromyalgia remains primarily idiopathic, case studies suggest that certain infections such as human immunodeficiency virus (HIV), hepatitis C virus, and Lyme disease may contribute to the development and/or worsening of fibromyalgia symptoms in some patients.^52^ Studies of chronic infections (HIV and Hepatitis C) have demonstrated increased rates of spondyloarthritis-spectrum disease and fibromyalgia symptoms.^53–55^ Likewise, infections have been implicated as potential triggers of disease activity in ERA and SpA; patients with these conditions display high rates of pain, fatigue, and sensory disturbances.^56–58^ We find it notable that these subtypes of arthritis (ERA and SpA) make up an overwhelming majority of the cases of arthritis observed in PANS.^24^ These findings could indicate shared post-infectious neuroimmune mechanisms.

The symptom patterns observed in this study closely resemble those reported in children and adults with post-acute sequelae of SARS-CoV-2 infection (“Long COVID”), including chronic fatigue, musculoskeletal pain, cognitive issues (“brain fog”), and post-exertional malaise.^59–61^ While a small subset of patients (n=7) had documentation of pre-deterioration COVID-19 infection in the EMR, we do not have systematic virologic testing to confirm causality or timing. Nevertheless, the overlap in symptom profiles suggests that convergent neuroimmune or post-infectious mechanisms may contribute to fibromyalgia symptoms in PANS, Long COVID, and other post-infectious conditions. Recognizing these shared features may help guide research towards unified models of post-infectious neuroimmune diseases and inform therapeutic strategies.

### Study Limitations

Our study is subject to several limitations. First, the assessment of our primary outcome relied on proxy-reported data. The components of the ACR FSQ (WPI and SSS) were often completed by caregivers. Although caregivers were instructed to complete questionnaires with the patient, patient participation was sometimes precluded by time constraints or severe neuropsychiatric symptoms. This creates the potential for discrepancies between a caregiver’s report and the patient’s direct experience, which may explain cases where clinicians suspected a pain syndrome before survey scores met the diagnostic threshold. Second, data collection was incomplete; ACR FSQ scores (WPI and SSS) were not systematically gathered during the clinic’s first years of operation, potentially resulting in the non-capture of early fibromyalgia cases. Only 36 of 204 patients had missing ACR FSQ or sleep disturbance data at presentation; because all 36 of these patients provided data at subsequent visits, the impact of this missing baseline information is likely minimal. The ACR FSQ has been validated for adolescent females, but it has not yet been evaluated in adolescent males or school-age children.^62^ Lastly, data collection occurred at discrete time points corresponding to clinic visits. As a result, the documented onset of fibromyalgia is later than the actual onset for most patients since symptoms often emerge between clinic visits.

### Clinical implications

Clinically, our findings emphasize the importance of proactive pain and fatigue screening in the evaluation and monitoring of PANS patients. Early intervention consists of a multi-pronged approach which may include graded exercise (aerobic and muscle strengthening), sleep hygiene coaching, cognitive behavioral therapy, acceptance and commitment therapy, mind-body exercises (qigong, tai chi, and yoga), adjunctive/complementary therapies (i.e., electroacupuncture), and pharmacological therapies for a subset that due not improve (tricyclics, gabapentinoids, SNRIs).^38,63–69^

### Conclusions

This study provides evidence that fibromyalgia symptoms can be long-term consequences for some patients with PANS. Notably, debilitating symptoms such as pain, fatigue, and cognitive symptoms (“brain fog”) often persisted even after psychiatric improvement, pointing to a potential shared pathophysiology between the two conditions. Further research is crucial to understand which patients are at risk for this chronic evolution and to develop targeted treatment strategies.

## Supporting information

Supplementary Online Content

## Data Availability

Deidentified participant data will be shared on reasonable request from any qualified investigator via email to.

## Author Contributions

Dr. Frankovich had full access to the data in the study and takes responsibility for the integrity of the data and the accuracy of the data analysis. Ms. Miles and Ms. Schlenk are co-first authors.

*Conceptualization:* Miles, Schlenk, Frankovich.

*Data curation*: Miles, Schlenk, Knight, Manko.

*Formal analysis:* Miles, Schlenk, Leonard, Challa, Gunther.

*Funding acquisition:* Frankovich.

*Investigation:* Miles, Ma, Farhadian, Gao, Manko, Xie, Tran, Silverman, Hoffman, Thienemann, Frankovich.

*Methodology:* Miles, Schlenk, Leonard, Challa, Gunther, Frankovich.

*Project administration:* Miles, Schlenk, Frankovich.

*Resources:* Ma, Farhadian, Manko, Xie, Tran, Silverman, Hoffman, Thienemann, Frankovich.

*Software:* Schlenk, Gunther.

*Supervision:* Frankovich.

*Validation:* Miles, Schlenk.

*Visualization:* Miles, Schlenk, Leonard, Challa, Gunther.

*Writing – original draft:* Miles, Schlenk, Gunther, Frankovich.

*Writing – review & editing:* Miles, Schlenk, Ma, Farhadian, Leonard, Knight, Challa, Gao, Gunther, Manko, Xie, Tran, Silverman, Hoffman, Thienemann, Frankovich.

## Acknowledgements

Funding for the infrastructure of our research program, training, advocacy, and overlapping projects came from: 1) Lucile Packard Foundation for Children’s Health; 2) National Institute of Mental Health-Pediatrics and Developmental Neuroscience Branch which supported the initial creation of the Stanford University IBH Research Program; 3) The Neuroimmune Foundation for education/training, microbial sequencing, and medication analyses; 4) The Brain Foundation and O’Sullivan Foundation for Autism research; 5) The Tara & Dave Dollinger PANS Biomarker Discovery Core for patient sample collection; 6) Stanford Maternal and Child Health Research Institute (MCHRI) for HLA research; 7) Stanford SPARK, SPARK Pisa, and International OCD Foundation for immunophenotyping research; 8) Oxnard Foundation for imaging research; 9) Caudwell Children’s Foundation for genetics and immunology research; 10) The Louisa Adelynn Johnson Fund for Complex Disease for expanding the Illuminate PANS project; 11) PPN for funding our healthy control project; 12) Stanford COMET, DRIVE, and HBREX programs for student research in our program; 13) LPCH Volunteer Program. We also wish to thank our dedicated clinic staff, collaborating clinicians, former members of our research staff who contributed to an early version of this project (Collin Leobold, Mattew Sigurdson, Avis Chan, Kayla Brown, Shirely Chen, Theresa Willet, Batuhan Bayram, and Kristin Sainani), and the many additional donors who have contributed to our work. Most importantly, we extend our deepest gratitude to the patients, families, and healthy control volunteers; this research would not be possible without their trust and participation.

