## Supplementary Online Content for "Prevalence and Predictors of Pain and Fatigue in Pediatric Acute-Onset Neuropsychiatric Syndrome (PANS)"

**eFigure 1.** Cohort Selection

**eAppendix 1.** Immune Behavioral Health (IBH) Screening Protocol Clinic Entry Screening Protocol

**eAppendix 2.** Healthy Controls

**eAppendix 3.** 2016 American College of Rheumatology (ACR) Fibromyalgia Criteria, Component Definitions, and Threshold Scores

**eAppendix 4.** Specifications of Data Capture System Used in This Study

**eAppendix 5.** Classification Criteria for Arthritis and Other Comorbid Autoimmune or Immunologic Conditions

**eTable 1.** Neuropsychiatric Symptoms and Scores at Clinic Presentation Among Consecutive Patients with PANS (N=204)

**eTable 2.** Characteristics of Pain, Fatigue, Cognitive, Sleep, and Sensory Symptoms Reported Any Time Within the First Year of Follow-Up in Consecutive Patients with PANS (N=204)

**eTable 3.** Severity of Pain and Sleep Disturbance Reported at One-Year Follow-Up Visit in Consecutive Patients with PANS (N=204) and at First Visit in Controls (N=111)

**eTable 4.** Characteristics of Pain, Fatigue, Cognitive, Sleep, and Sensory Symptoms Among Patients with PANS at Time Point Meeting American College of Rheumatology (ACR) Fibromyalgia Criteria Threshold Scores (N=85)

**eTable 5.** Clinical Characteristics of Consecutive Patients with PANS, Stratified by Satisfaction of American College of Rheumatology (ACR) Fibromyalgia Criteria Threshold Scores and Progression to Full ACR Fibromyalgia Criteria (N=204)

**eTable 6.** Cumulative Incidence of Meeting American College of Rheumatology (ACR) Fibromyalgia Criteria Threshold Scores and Full ACR Fibromyalgia Criteria Among Consecutive Patients with PANS for Different Time Points by Years After Presentation to Clinic (N=204)

**eTable 7.** Cumulative Incidence of Meeting American College of Rheumatology (ACR) Fibromyalgia Criteria Threshold Scores and Full ACR Fibromyalgia Criteria Among Consecutive Patients with PANS for Different Time Points by Age (N=204)

**eAppendix 6.** Pain Syndrome Documentation

**eFigure 3.** Longitudinal Trajectories of Widespread Pain Index (WPI) Scores in Consecutive Patients with PANS (N=204)

**eFigure 4.** Longitudinal Trajectories of Waking Unrefreshed in Consecutive Patients with PANS (N=204), Part 1 of 3

**eFigure 5.** Longitudinal Trajectories of Daytime Fatigue in Consecutive Patients with PANS (N=204), Part 2 of 3

**eFigure 6.** Longitudinal Trajectories of Cognitive Symptoms (“Brain Fog”) in Consecutive Patients with PANS (N=204), Part 3 of 3

**eTable 8.** Exploratory Univariable Logistic Regression of Candidate Baseline Covariates

**eTable 9.** Correlations and Variance Inflation Factors Among Covariates Included in Exploratory Cox Proportional Hazard Model

**eFigure 1. Cohort Selection**

679 patients evaluated by the Immune Behavioral Health (IBH) Clinic

Exclusions:

- Not a local patient^c^ (n=22)
- <3 visits (n=42)
- <4y at presentation (n=5)^d^
- ≥18y at presentation (n=9)^d^
- <9mo of follow-up (n=31)
- Non-index sibling^e^ (n=17)

330 patients with PANS consented for research^b^

204 patients included in study

373 patients with PANS

43 patients with PANS not enrolled in research

~6,800 patients screened by the Immune Behavioral Health (IBH) Clinic between 15 September 2012 and 1 January 2026^a^

303 patients without PANS

36 patients missing symptom scores at intake^f^

168 patients included in Cox proportional hazard analysis^f^

^a^Priority was given to local patients and patients with high likelihood of meeting PANS criteria based on screening form, electronic record review, +/- discussion with referring provider, +/- discussion with primary doctor*.* IBH screening protocol is described in **eAppendix 1.**

^b^Patients are considered eligible for this study if they have provided consent/assent for our prospective biomarker study (IRB#26922).

^c^The IBH Clinic is located in Menlo Park, California, United States. For this study, patients were considered local if they primarily live within the following regions (counties) at clinical presentation: Superior California (Butte, Colusa, El Dorado, Glenn, Lassen, Modoc, Nevada, Placer, Plumas, Sacramento, Shasta, Sierra, Siskiyou, Sutter, Tehama, Yolo, Yuba), North Coast (Del Norte, Humboldt, Lake, Mendocino, Napa, Sonoma, Trinity), San Francisco Bay Area (Alameda, Contra Costa, Marin, San Francisco, San Mateo, Santa Clara, Solano), Northern San Joaquin Valley (Alpine, Amador, Calaveras, Madera, Mariposa, Merced, Mono, San Joaquin, Stanislaus, Tuolumne), or Central Coast (Monterey, San Benito, San Luis Obispo, Santa Barbra, Santa Cruz, Ventura).

^d^All patients included in this study were 18 years or younger at clinic presentation.

^e^In circumstances where two or more siblings are considered for study inclusion, we elected to include only the first sibling to present to the IBH Clinic (also called the index sibling). This individual is commonly the more severe case in their respective sibling group.

^f^The Fibromyalgia Survey Questionnaire (FSQ) (which includes the Widespread Pain Index [WPI] and Symptom Severity Scale [SSS]) and other measures were on paper and each item was optional until 2017. Nineteen patients evaluated before 2017 are missing these values and are excluded from our Cox proportional hazard model. An additional 17 patients are missing sleep disturbance data and are also excluded from the model.

*Abbreviations*: Pediatric acute-onset neuropsychiatric syndrome (PANS); Immune Behavioral Health (IBH).

**eAppendix 1. Immune Behavioral Health (IBH) Screening Protocol Clinic Entry Screening Protocol**

To ethically manage an overwhelming number of patient referrals that had become unsustainable, we sought guidance from the Stanford Ethics Committee. In consultation with them, we established a standardized screening protocol in September 2014, which is administered by a single nurse practitioner (BF). This protocol provides a fair and consistent method for managing clinic capacity and fulfilling our research mission by prioritizing local patients and individuals who facilitate research on early, untreated disease flares.

Priority is given to patients who meet the following criteria:

**1. Geographic Location:** The patient is a resident of one of 13 specified counties in Northern California surrounding the clinic location: Alameda, Contra Costa, San Francisco (surveyed by CDC), Marin, San Mateo, Santa Clara, Santa Cruz, Monterey, Napa, Sonoma, Solano, San Benito, Sacramento.

**2. Referral Source:** The patient has a referral from a board-certified pediatrician, family practice physician, developmental pediatrician, neurologist, psychiatrist, psychologist, or equivalent.

**3. Medical Records:** A minimum of three years of medical records preceding the symptomatic deterioration are available during the screening process. If the patient is accepted, all medical records from birth to the present are required.

**4. Primary Medical Doctor (PMD) Relationship:** The patient has an established relationship with a local primary medical doctor who has clearly documented a stable functional baseline prior to the deterioration. Preference is given to patients who were followed by the same clinic for at least 3 years prior to the deterioration so that the prior baseline is known to the primary practice.

**5. Record Review Clarification:** In cases where a review of records is inconclusive, the PMD is available for a case consultation with the screening nurse practitioner to assess the likelihood of pediatric acute-onset neuropsychiatric syndrome (PANS).

**6. Comorbid Conditions:** Preexisting, stable diagnoses such as autism spectrum disorder (ASD), attention-deficit/hyperactivity disorder (ADHD), obsessive-compulsive disorder (OCD), or an anxiety disorder are permissible, provided the patient demonstrates stable baseline functioning (e.g., attending school) prior to the sudden onset of new psychiatric/behavioral symptoms.

**7. Symptom Onset:** The symptom onset is hyperacute (a transition from a stable baseline to severe symptoms in less than 72 hours) or acute (a transition from a stable baseline to severe symptoms within 3-7 days).

**8. Clinical Course and Timing:** Preference is given to patients with a single episode or a relapsing-remitting course who are in a flare for less than four months. As clinic capacity permits, patients up to nine months from the start of a flare are considered.

**9. Diagnostic Probability:** Based on a record review and potential discussion with the PMD, there is a high likelihood that the patient fulfills PANS criteria. The PMD, referring clinician(s), and the screening nurse practitioner must all have a strong clinical suspicion for PANS.

**10. Prior Treatment Status:** The patient is relatively untreated. The following prior treatments are permitted: a course of antibiotics for an infection (e.g., streptococcal pharyngitis) and standard-of-care medications for psychiatric symptoms.

**11. Vaccination Status:** The patient has completed all age-appropriate vaccinations as recommended by the Centers for Disease Control and Prevention (CDC) and the American Academy of Pediatrics (AAP). This prerequisite mitigates risks associated with possible immunomodulatory therapies and prevents exposure to other immunocompromised patients in the shared waiting room.

**Notes on Protocol Implementation:**

**Flexibility:** During periods of low referral volume (such as summer months or the 2020-2021 quarantine period), the protocol allows for the inclusion of patients who are missing one or two of the above criteria.

**Education:** To improve the referral pipeline, the program provides educational outreach on PANS to local PMDs via grand rounds, lectures, and telephone consultations.

**Protocol History:** This standardized screening protocol has been in effect since September 2014. Between September 2012 and September 2014, all referred patients were accepted into the clinic. Prior to 15 June 2022, the geographic location criterion was restricted to 8 surrounding counties.

**eAppendix 2. Healthy Controls**

Controls are recruited from the following nine Northern California counties which represent the majority of our patient base: Alameda, Contra Costa, Marin, Napa, San Francisco, San Mateo, Santa Clara, Solano, and Sonoma. Recruitment materials include: 1) Physical flyers (distributed to community centers, Girls and Boys Club, libraries, dentist offices, schools, etc.) and 2) postcards, and Epic message distributed to qualifying individuals via Stanford Honest Broker Research Program. Recruitment materials are accessible online via Studypages and Idealist. Controls and/or their caregivers fill out a screening form accessed through a QR code on research outreach materials. Upon passing the initial screen, controls and/or their caregivers are invited to provide formal consent for the participation in research. After the consent process is finalized, controls and/or their caregivers are asked to complete the Stanford Immune Behavioral Health (IBH) questionnaire, which mirrors the questionnaire distributed to patients. Questionnaire respondents were encouraged to ask the research assistant questions to avoid errors.

Controls were included in the current study if they had at least one questionnaire with completed American College of Rheumatology (ACR) Fibromyalgia Survey Questionnaire (FSQ) from before age 18. Among sets of healthy control siblings considered for this study, we selected only the first sibling with a complete data set. We included controls who had first degree family members with and without a family history of autoimmune, immunologic, or psychiatric disease.

**eAppendix 3. 2016 American College of Rheumatology (ACR) Fibromyalgia Criteria, Component Definitions, and Threshold Scores**

| **Component** | **Definition** |
| --- | --- |
| *Widespread Pain Index (WPI)* | The WPI is a patient- or caregiver- reported count of the number of specific areas of the body (out of 19) with pain present over the past week. Respondents are asked to not report pain due to injury.  The areas include left and right shoulder, left and right upper arm, left and right lower arm, left and right hip, left and right upper leg, left and right lower leg, jaw (left or right), chest, abdomen, upper back, lower back, and neck.  The final score is calculated from the sum of all responses (min 0, max 19). |
| *Symptom Severity Scale (SSS)* | The SSS is comprised of two components:   1. Three core symptoms (**daytime fatigue, waking unrefreshed,** and **cognitive symptoms**) rated as none (0), mild (1), moderate (2), or severe (3) in the past week. Points are added to the sum based on the level of severity of each symptom. 2. A checklist of somatic symptoms in the past six months or since last visit (each worth one point in score calculation): **headaches, abdominal pain/cramps,** and **depression**.   The final score is calculated from the sum of all responses (min 0, max 12). |
| *Generalized Pain* | Generalized pain is defined as pain in at least 4 of 5 regions. Regions of pain are based on the responses in the WPI as shown below. The sum of regions excludes pain in the jaw, chest, and abdomen.   - Region 1 (left upper): Left shoulder, upper arm, lower arm - Region 2 (right upper): Right shoulder, upper arm, lower arm - Region 3 (left lower): Left hip, upper leg, lower leg - Region 4 (right lower): Right hip, upper leg, lower leg - Region 5 (axial): upper back, lower back, neck |
| **Full Fibromyalgia Criteria**^2^ | **Full ACR fibromyalgia criteria require that all three are met**:   1. Generalized pain, defined as pain in at least 4 of 5 regions, is present. 2. Symptoms have been present at a similar level for at least 3 months. 3. WPI ≥ 7 and SSS ≥ 5 OR WPI ≥ 4 and SSS ≥ 9 (***threshold scores***) |

The WPI and SSS together make up the Fibromyalgia Survey Questionnaire (FSQ).

*Abbreviations*: American College of Rheumatology (ACR); Widespread Pain Index (WPI); Symptom Severity Scale (SSS); Fibromyalgia Survey Questionnaire (FSQ).

**eAppendix 4. Specifications of Data Capture Used in This Study**

The Stanford REDCap platform is developed and operated by Stanford Medicine Research Technology team. The REDCap platform services at Stanford are subsidized by a) Stanford School of Medicine Research Office, and b) the National Center for Research Resources and the National Center for Advancing Translational Sciences, National Institutes of Health, through grant UL1 TR003142.

Patient surveys were given using paper format prior to 2017 and while families were asked to complete the entire paper questionnaire, occasionally items would be missed. Data from the paper forms were entered into a REDCap database. After 2017, patient surveys were administered via REDCap.

**eAppendix 5. Classification Criteria for Juvenile Arthritis and Other Comorbid Autoimmune or Immunologic Conditions**

| **Condition** | **Criteria** |
| --- | --- |
| Juvenile Idiopathic Arthritis (JIA) (ACR) | Must be diagnosed by a pediatric rheumatologist using the following criteria:  1. Arthritis onset **before age 16** and  2. Arthritis in **one or more joints as defined as A or B**:  A. Swelling/effusion(s) (documented by physical exam, ultrasound, or MRI),  B. One of: limited motion, pain with motion, tenderness, or warmth,  and  3. Arthritis duration of **at least six weeks** in the same joint, and 4. Other causes of arthritis excluded |
| **Autoimmune Conditions** | |
| Celiac Disease | Must be diagnosed by a pediatric gastroenterologist and documented in the EMR. Must have either biopsy-proven disease or high-risk serotype (HLA-DQ2 or HLA-DQ8) coupled with high tissue transglutaminase antibodies. |
| Psoriasis | Must be diagnosed by a dermatologist and documented in the EMR. |
| Thyroiditis | Must be diagnosed by a pediatrician or pediatric endocrinologist (documented in the EMR) and have at least one of the following:   1. An ultrasound showing thyroid fullness or 2. Elevated thyroid antibodies (TPO or anti-thyroglobulin) on two separate occasions |
| Inflammatory bowel disease (IBD) | Must be diagnosed by a pediatric gastroenterologist (documented in the EMR) and have endoscopic evidence of IBD. |
| Systemic lupus erythematosus (SLE) | Must be classified as SLE by a pediatric rheumatologist using the EULAR/ACR classification criteria for SLE: Patient must have a positive ANA (≥ 1:80, entry criterion) with at least one clinical criterion and ≥ 10 points accumulated across multiple domains. Each clinical (constitutional, hematologic, neuropsychiatric, mucocutaneous, serosal, musculoskeletal, renal) and immunologic (antiphospholipid antibodies, complement proteins, SLE specific antibodies) domain contain a weighted number. The highest weighted criterion in each domain is counted toward the total score. Occurrence of a criterion on at least one occasion is sufficient. Criteria need not occur simultaneously. |
| **Other Immunologic Conditions^a^** | |
| Eosinophilic Esophagitis (EoE) | Must be diagnosed by a pediatric allergist or gastroenterologist (documented in the EMR) using the following criteria:   1. Symptoms of esophageal dysfunction, or concomitant atopic conditions, or endoscopic findings of rings, furrows, exudates, edema, stricture, narrowing, and crepe-paper mucosa, and 2. Greater than or equal to 15 eos/hpf (~60 eos/mm^2^) on esophageal biopsy, and   Exclusion of non-EoE disorders that cause or potentially contribute to EoE. |
| Primary Immunodeficiency (PID) | Must be diagnosed by a pediatric immunologist and documented in the EMR.   1. Common variable immunodeficiency (CVID): Patient has markedly reduced serum IgG levels in combination with low levels of IgA and/or IgM (<2 SD below the mean), poor or absent response to immunizations, and an absence of any other defined immunodeficiency state. 2. Specific polysaccharide antibody deficiency (SPAD): Patient has a poor response to Pneumovax, i.e., <70% of 23 protective serotypes in children 6 years or older, or <50% in children younger than 6 years. 3. Hypogammaglobulinemia: Patient has a reduction in any serum immunoglobulin levels (IgG, IgA, or IgM).   Selective IgA deficiency: Patient older than 4 years has an isolated deficiency of IgA (i.e., in the setting of normal levels of IgG and IgM) and other causes of hypogammaglobulinemia have been excluded |
| Chronic Urticaria | Must be diagnosed by a pediatrician or pediatric immunologist, or symptoms must have required prolonged steroid treatment (6 or more weeks). |

^a^We do not include “other immunologic conditions” In **Figure 2.**

We did not include diagnoses made by non-board-eligible clinicians (i.e., clinicians who did not complete training at a program accredited by Accreditation Council for Graduate Medical Education [ACGME] or the Royal College of Physicians and Surgeons of Canada [RCPSC]).

*Abbreviations*: Juvenile Idiopathic Arthritis (JIA); American College of Rheumatology (ACR); Electronic medical record (EMR); Human leukocyte antigen (HLA); Thyroid peroxidase (TPO); Inflammatory bowel disease (IBD); Systemic lupus erythematosus (SLE); American College of Rheumatology (ACR); European Alliance of Associations for Rheumatology (EULAR); Anti-nuclear antibody (ANA); Eosinophilic esophagitis (EoE); Eosinophils (eos); High-powered field (hpf); Primary Immunodeficiency (PID); Common variable immunodeficiency (CVID); Immunoglobulin G (IgG); Immunoglobulin A (IgA); Specific polysaccharide antibody deficiency (SPAD); Immunoglobulin M (IgM).

**eTable 1. Neuropsychiatric Symptoms and Scores at Clinic Presentation Among Consecutive Patients with PANS (N=204)**

| **Characteristic** | **Patients (N=204)** |
| --- | --- |
| **Symptoms^a^ at Clinic Presentation, No. (%)** | |
| Obsessive-compulsive symptoms | 185 (90.7) |
| Food/fluid refusal or restriction | 108 (52.9) |
| Urge to overeat | 20 (9.8) |
| Separation anxiety | 126 (61.8) |
| Other anxiety (fears, phobias, panic) | 179 (87.7) |
| Depression with or without suicidal ideation | 120 (58.8) |
| Moodiness or mood swings | 146 (71.6) |
| Emotional lability (inappropriate crying or laughing spells) | 82 (40.2) |
| Irritability, aggression, and/or oppositional behaviors | 167 (81.9) |
| Hyperactivity or trouble paying attention | 86 (42.2) |
| Behavioral regression (baby talk, poor self-care, immature judgement) | 99 (48.5) |
| Cognitive symptoms (difficulty thinking, “brain fog,” memory problems) | 112 (54.9) |
| Worsening of school performance | 102 (50.0) |
| Dysgraphia | 62 (30.4) |
| Sensory amplification | 122 (59.8) |
| Sleep disturbance | 141 (69.1) |
| Urinary symptoms (enuresis or increased urinary frequency) | 86 (42.2) |
| Psychotic symptoms (hallucinations, delusions, paranoia) | 59 (28.9) |
| Tics (motor or phonic) | 100 (49.0) |
| No. symptoms at presentation (above), mean (SD) | 10.3 (3.6) |
| **Psychometric Scores at Clinical Presentation^b^** | |
| Global Impairment, mean (SD), n=185 | 47.8 (24.1) |
| Caregiver Burden Inventory, mean (SD), n=152 | 34.5 (17.9) |
| Widespread Pain Index, mean (SD), n=185 | 1.9 (2.4) |
| Malaise Score (Symptom Severity Score), mean (SD), n=183 | 4.9 (2.9) |
| Children’s Global Assessment Scale (Clinician Rating), mean (SD), n=145 | 58.4 (16.6) |

^a^Symptoms presented are from the 31-Item PANS Scale.^28^

^b^Data reported are the first captured scores per patient within 90 days of presentation to clinic.

*Abbreviations*: Pediatric acute-onset neuropsychiatric syndrome (PANS); Immune Behavioral Health (IBH); standard deviation (SD).

**eTable 2. Characteristics of Pain, Fatigue, Cognitive, Sleep, and Sensory Symptoms Reported Any Time Within the First Year of Follow-Up in Consecutive Patients with PANS (N=199)**

| **Characteristic** | **Patients / No. Patients with Available Data from First Year of Follow-up (N=199) (%)** |
| --- | --- |
| **ACR FSQ: Widespread Pain Index (WPI)^a^ and Pain Regions, No. (%)** | |
| Any pain in the past week (WPI > 0) | 180 (90.5) |
| Body part with pain | |
| Jaw | 53 (26.6) |
| Neck | 96 (48.2) |
| Chest | 59 (29.6) |
| Abdomen | 126 (63.3) |
| Back (upper or lower) | 100 (50.3) |
| Shoulder | 67 (33.7) |
| Hip | 59 (29.6) |
| Arm (upper or lower) | 70 (35.2) |
| Leg (upper or lower) | 104 (52.3) |
| WPI ≥ 4 and WPI < 7 | 82 (41.2) |
| WPI ≥ 7 | 40 (20.1) |
| Four or more regions of pain^a^ at one time | 44 (22.1) |
| **ACR FSQ: Symptom Severity Scale (SSS), No. (%)** | |
| Moderate or severe symptoms in the past week | |
| Daytime fatigue | 136 (68.3) |
| Waking unrefreshed | 129 (64.8) |
| Cognitive symptoms (“brain fog”) | 113 (56.8) |
| Symptoms present in past 6 months | |
| Headaches | 156 (78.4) |
| Pain/cramps in abdomen | 151 (75.9) |
| Depression | 134 (67.3) |
| SSS ≥ 5 and SSS < 9 | 144 (72.4) |
| SSS ≥ 9 | 59 (29.6) |
| **Other Symptoms Endorsed on the Stanford University IBH Clinic Questionnaire, No. (%)** | |
| Sleep disturbance^b^ | 154 (77.4)^b^ |
| Exercise intolerance^c^ | 68 (34.2)^c^ |
| Sensory disturbances (any below) | 163 (81.9) |
| Shooting or burning pain | 36 (18.1) |
| Numbness | 32 (16.1) |
| Temperature intolerance (any below)^c^ | 50 (25.1)^c^ |
| Cold intolerance^c^ | 34 (17.1)^c^ |
| Heat intolerance^c^ | 40 (20.1)^c^ |
| Sensory amplification (any below) | 160 (80.4) |
| Noise sensitivity | 115 (57.8) |
| Light sensitivity | 83 (41.7) |
| Smell/taste sensitivity | 84 (42.2) |
| Touch/texture sensitivity^c^ | 33 (16.6)^c^ |
| Sensory amplification (non-specific) | 74 (37.2) |
| **Fibromyalgia Criteria Threshold Scores**^d^ **and Full Criteria**^e^ **Any Time Within First Year, No. (%)** | |
| Meets ACR fibromyalgia criteria threshold scores^d,f^ | 46 (24.1)^f^ |
| Meets full ACR fibromyalgia criteria^e,f^ | 9 (4.7)^f^ |

^a^The WPI does not include pain attributable to injury. The sum of WPI regions excludes pain in the chest, abdomen, and jaw.

^b^This symptom was not a required question in the IBH patient questionnaire until 2017. Percentages are calculated from the number of patients (n=191 of 199) for whom data is available from within the first year of follow-up.

^c^This symptom was not part of the IBH patient questionnaire until 2017. Percentages are calculated from the number of patients (n=155 of 199) for whom one or more questionnaires from the first year of follow-up captures the symptom.

^d^ACR fibromyalgia criteria threshold scores (WPI ≥ 7 and SSS ≥ 5 OR WPI ≥ 4 and SSS ≥ 9) are derived from the ACR FSQ and based on 2016 ACR fibromyalgia criteria (**eAppendix 3**).

^e^To meet full ACR fibromyalgia criteria, patient must meet aforementioned threshold scores and have pain in 4 of 5 body regions (using the WPI) and pain must persist for 3 or more months.

^f^Percentages calculated out of total number of patients with FSQ (WPI and SSS) scores available in the first year of follow-up (n=191 of 199 patients).

*Abbreviations*: Pediatric acute-onset neuropsychiatric syndrome (PANS); American College of Rheumatology (ACR); Widespread Pain Index (WPI); Symptom Severity Scale (SSS); Immune Behavioral Health (IBH); Fibromyalgia Survey Questionnaire (FSQ).

**eTable 3. Severity of Pain and Sleep Disturbance Reported at One-Year Follow-Up Visit in Consecutive Patients with PANS (N=204) and at First Visit in Controls (N=111)**

| **Symptom** | **PANS (N=204)** | **Controls (N=111)** |
| --- | --- | --- |
| **Pain^a^, no. / total no. with severity rating (%)** | | |
| None | 50/92 (54.3) | 102/111 (91.9) |
| Mild | 23/92 (25.0) | 8/111 (7.2) |
| Moderate | 13/92 (14.1) | 1/111 (0.9) |
| Severe | 5/92 (5.4) | 0 |
| Extreme | 1/92 (1.1) | 0 |
| **Sleep Disturbance^a^, no. / total no. with severity rating (%)** | | |
| None | 72/117 (61.5) | 96/111 (86.5) |
| Mild | 23/117 (19.7) | 15/111 (13.5) |
| Moderate | 14/117 (12.0) | 0 |
| Severe | 6/117 (5.1) | 0 |
| Extreme | 2/117 (1.7) | 0 |

*^a^Pain (headaches, abdominal pain, body pain)* and *Sleep disturbance* are part of the 31-Item PANS Scale, which is administered at every patient visit after 2017.^28^ Earlier versions of the scale recorded *Sleep disturbance* as a dichotomous (yes/no) item. We report the distribution of these data at the time of the one-year follow-up visit among patients for whom a severity score was available. We additionally report symptom severities among controls.

**eTable 4. Characteristics of Pain, Fatigue, Cognitive, Sleep, and Sensory Symptoms Among Patients with PANS at Time Point Meeting American College of Rheumatology (ACR) Fibromyalgia Criteria Threshold Scores (N=85)**

| **Characteristic** | **Met Fibromyalgia Criteria Threshold Scores (WPI and SSS)^a^** *Without Progression to Full ACR Fibromyalgia* | **Met Full ACR Fibromyalgia Criteria^b^** |
| --- | --- | --- |
| **No. Patients** | **57** | **28** |
| **ACR FSQ: Widespread Pain Index (WPI)^c^ and Pain Regions** | | |
| Any pain in the past week (WPI > 0), No. (%) | 57 (100.0) | 28 (100.0) |
| Body part with pain, No. (%) | | |
| Jaw | 6 (10.5) | 9 (32.1) |
| Neck | 31 (54.4) | 18 (64.3) |
| Chest | 17 (29.8) | 8 (28.6) |
| Abdomen | 27 (47.4) | 16 (57.1) |
| Back (upper or lower) | 42 (73.7) | 20 (71.4) |
| Shoulder | 24 (42.1) | 18 (64.3) |
| Hip | 20 (35.1) | 13 (46.4) |
| Arm (upper or lower) | 23 (40.4) | 15 (53.6) |
| Leg (upper or lower) | 41 (71.9) | 24 (85.7) |
| WPI ≥ 4 and WPI < 7, No. (%) | 22 (38.6) | 9 (32.1) |
| WPI ≥ 7, No. (%) | 35 (61.4) | 19 (67.9) |
| WPI, mean (SD) | 7.3 (2.7) | 8.7 (3.9) |
| Four of more regions of pain^c^ at one time, No. (%) | 25 (43.9) | 16 (57.1) |
| **ACR FSQ: Symptom Severity Scale (SSS)** | | |
| Moderate or severe symptoms in the past week, No. (%) | | |
| Daytime fatigue | 42 (73.7) | 26 (92.9) |
| Waking unrefreshed | 40 (70.2) | 22 (78.6) |
| Cognitive symptoms (“brain fog”) | 34 (59.6) | 16 (57.1) |
| Symptoms present in past 6 months, No. (%) | | |
| Headaches | 43 (75.4) | 27 (96.4) |
| Pain/cramps in abdomen | 39 (68.4) | 25 (89.3) |
| Depression | 40 (70.2) | 21 (75.0) |
| SSS ≥ 5 and SSS < 9, No. (%) | 29 (50.9) | 10 (35.7) |
| SSS ≥ 9, No. (%) | 28 (49.1) | 18 (64.3) |
| SSS, mean (SD) | 7.9 (2.1) | 8.8 (2.0) |
| **Other Symptoms Endorsed on the Stanford University IBH Clinic Questionnaire, No. (%)** | | |
| Sleep disturbance^d^ | 38 (73.1)^d^ | 15 (75.0)^d^ |
| Exercise intolerance^e^ | 18 (36.0)^e^ | 4 (28.6)^e^ |
| Sensory disturbances (any below) | 37 (64.9) | 20 (71.4) |
| Shooting or burning pain | 9 (15.8) | 6 (21.4) |
| Numbness | 1 (1.8) | 3 (10.7) |
| Temperature intolerance (any below)^e^ | 6 (12.0)^e^ | 4 (28.6)^e^ |
| Cold intolerance^e^ | 5 (10.0)^e^ | 4 (28.6)^e^ |
| Heat intolerance^e^ | 1 (2.0)^e^ | 1 (7.1)^e^ |
| Sensory amplification (any below) | 37 (64.9) | 20 (71.4) |
| Noise sensitivity | 21 (36.8) | 14 (50.0) |
| Light sensitivity | 13 (22.8) | 8 (28.6) |
| Smell/taste sensitivity | 11 (19.3) | 9 (32.1) |
| Touch/texture sensitivity^e^ | 7 (14.0)^e^ | 0^e^ |
| Sensory amplification (non-specific) | 5 (8.8) | 2 (7.1) |

^a^ ACR fibromyalgia criteria threshold scores (WPI ≥ 7 and SSS ≥ 5 OR WPI ≥ 4 and SSS ≥ 9) are derived from the ACR FSQ and based on 2016 ACR fibromyalgia criteria (**eAppendix 3**).

^b^To meet full ACR fibromyalgia criteria, patient must meet aforementioned threshold scores and have pain in 4 of 5 body regions (using the WPI) and pain must persist for 3 or more months.

^c^The WPI does not include pain attributable to injury. The sum of WPI regions excludes pain in the chest, abdomen, and jaw.

^d^This symptom was not a required question in the IBH patient questionnaire until 2017. Percentages are calculated from the number of patients (n=52, 20) for whom data is available.

^e^This symptom was not part of the IBH patient questionnaire until 2017. Percentages are calculated from the number of patients (n=50, 14) whose questionnaire captured the symptom.

*Abbreviations*: Pediatric acute-onset neuropsychiatric syndrome (PANS); American College of Rheumatology (ACR); Widespread Pain Index (WPI); Symptom Severity Scale (SSS); Fibromyalgia Survey Questionnaire (FSQ); standard deviation (SD); Immune Behavioral Health (IBH).

**eTable 5. Clinical Characteristics of Consecutive Patients with PANS, Stratified by Satisfaction of American College of Rheumatology (ACR) Fibromyalgia Criteria Threshold Scores and Progression to Full ACR Fibromyalgia Criteria (N=204)**

| **Characteristic** | **Never met Fibromyalgia Criteria Threshold Scores^a^** | **Met Fibromyalgia Criteria Threshold Scores (WPI and SSS)^a,b^**  *Without Progression to Full ACR Fibromyalgia* | **Met Full ACR Fibromyalgia Criteria^b^** |
| --- | --- | --- | --- |
| No. Patients | **119** | **57** | **28** |
| Male, No. (%) | 62 (52.1) | 31 (54.4) | 17 (60.7) |
| White or European, No. (%) | 90 (75.6) | 42 (73.7) | 19 (67.9) |
| Age at first-ever neuropsychiatric deterioration, mean (SD), years | 8.1 (3.3) | 8.4 (3.3) | 8.3 (3.5) |
| Age at deterioration prompting referral to clinic, mean (SD), years | 9.2 (3.3) | 10.0 (3.1) | 10.1 (2.9) |
| Age at PANS onset, mean (SD), years | 9.3 (3.4) | 9.4 (3.3) | 10.5 (3.4) |
| Age at IBH Clinic presentation, mean (SD), years | 9.7 (3.4) | 10.5 (3.3) | 11.2 (2.2) |
| Met PANS criteria at or before clinic presentation, No. (%) | 103 (86.6) | 53 (93.0) | 22 (78.6) |
| IBH Clinic follow-up time, mean (SD), years | 4.3 (2.9) | 6.0 (3.4) | 7.1 (3.1) |
| Clinical course at time of study, No. (%) | | | |
| Single flare | 15 (12.8) | 10 (17.9) | 11 (39.9) |
| Relapsing-remitting | 76 (65.0) | 30 (53.6) | 11 (39.9) |
| Primary persistent | 11 (9.4) | 2 (3.6) | 0 |
| Secondary persistent | 15 (12.8) | 14 (25.0) | 6 (21.4) |
| **Scores at Clinical Presentation^c^** | | | |
| Global Impairment, mean (SD) | 45.2 (24.7) | 52.5 (22.9) | 51.8 (20.9) |
| Caregiver Burden Inventory, mean (SD) | 29.9 (15.8) | 42.8 (18.4) | 42.6 (16.9) |
| Widespread Pain Index^d^, mean (SD) | 1.1 (1.5) | 2.8 (2.6) | 4.4 (3.2) |
| Symptom Severity Scale, mean (SD) | 3.8 (2.3) | 6.2 (2.8) | 7.1 (3.1) |
| Children’s Global Assessment Scale, mean (SD) | 61.4 (15.6) | 54.9 (18.7) | 47.0 (8.4) |
| **Comorbid Conditions^e^** | | | |
| Any chronic arthritis, No. (%) | 17 (14.3) | 20 (35.1) | 20 (71.4) |
| Any autoimmune disease other than arthritis, No. (%) | 7 (5.9) | 6 (10.5) | 8 (28.6) |
| Any other immunologic disease, No. (%) | 2 (1.7) | 3 (5.3) | 3 (10.7) |

^a^ ACR fibromyalgia criteria threshold scores (WPI ≥ 7 and SSS ≥ 5 OR WPI ≥ 4 and SSS ≥ 9) are derived from the ACR FSQ and based on 2016 ACR fibromyalgia criteria (**eAppendix 3**).

^b^To meet full ACR fibromyalgia criteria, patient must meet aforementioned threshold scores and have pain in 4 of 5 body regions (using the WPI) and pain must persist for 3 or more months.

^c^Data reported are the first captured scores per patient within 90 days of presentation to clinic. We report the total number of patients with available data per row (descending in order—Global Impairment, Caregiver Burden Inventory, Widespread Pain Index, Symptom Severity Scale, Children’s Global Assessment Scale): No fibromyalgia (n=112, 94, 113, 113, 93); meets ACR fibromyalgia criteria threshold scores only (n=51, 40, 52, 51, 41); meets full ACR fibromyalgia criteria (n=24, 20, 22, 21, 13).

^d^The WPI does not include pain attributable to injury. The sum of WPI regions excludes pain in the chest, abdomen, and jaw.

^e^See **eAppendix 5** for classification criteria of arthritis, autoimmune diseases and other immunologic conditions.

*Abbreviations*: Pediatric acute-onset neuropsychiatric syndrome (PANS); American College of Rheumatology (ACR); Widespread Pain Index (WPI); Symptom Severity Scale (SSS); Immune Behavioral Health (IBH); standard deviation (SD); Fibromyalgia Survey Questionnaire (FSQ).

**eTable 6. Cumulative Incidence of Meeting American College of Rheumatology (ACR) Fibromyalgia Criteria Threshold Scores and Full ACR Fibromyalgia Criteria Among Consecutive Patients with PANS for Different Time Points by Years After Presentation to Clinic (N=204)**

| **At time (years after presentation)** | **Time to ACR fibromyalgia criteria threshold scores among 204 patients with PANS** | | | | **Time to full ACR fibromyalgia criteria among 204 patients with PANS** | | | |
| --- | --- | --- | --- | --- | --- | --- | --- | --- |
|  | **No. at risk** | **No. censored** | **No. events** | **Cumulative incidence, % (95% CI)** | **No. at risk** | **No. censored** | **No. events** | **Cumulative incidence, % (95% CI)** |
| 0 | 204 | 0 | 17 | 8.3 (5.3–13.1) | 204 | 0 | 2 | 1.0 (0.2–3.9) |
| 1 | 149 | 9 | 46 | 22.6 (17.4–29.0) | 182 | 13 | 9 | 4.4 (2.3–8.3) |
| 2 | 106 | 35 | 63 | 32.6 (26.4–39.8) | 146 | 46 | 12 | 6.2 (3.6–10.7) |
| 3 | 83 | 50 | 71 | 38.1 (31.4–45.7) | 123 | 65 | 16 | 8.9 (5.5–14.2) |
| 4 | 65 | 64 | 75 | 41.4 (34.4–49.3) | 100 | 85 | 19 | 11.2 (7.2–17.2) |
| 5 | 56 | 73 | 75 | 41.4 (34.4–49.3) | 84 | 96 | 24 | 16.0 (10.8–23.4) |
| 6 | 40 | 85 | 79 | 45.9 (38.2–54.3) | 65 | 113 | 26 | 18.3 (12.6–26.2) |
| 7 | 26 | 96 | 82 | 50.6 (42.0–59.9) | 52 | 126 | 26 | 18.3 (12.6–26.2) |
| 8 | 18 | 104 | 82 | 50.6 (42.0–59.9) | 38 | 140 | 26 | 18.3 (12.6–26.2) |
| 9 | 12 | 110 | 82 | 50.6 (42.0–59.9) | 25 | 152 | 27 | 20.5 (13.9–29.7) |
| 10 | 8 | 112 | 84 | 60.5 (46.7–74.5) | 16 | 160 | 28 | 24.1 (15.6–36.2) |
| 11 | 1 | 118 | 85 | 65.4 (50.3–80.1) | 3 | 173 | 28 | 24.1 (15.6–36.2) |
| 12 | 0 | 119 | 85 | 65.4 (50.3–80.1) | 0 | 176 | 28 | 24.1 (15.6–36.2) |

*Abbreviations*: American College of Rheumatology (ACR); Pediatric acute-onset neuropsychiatric syndrome (PANS); Confidence interval (CI).

**eTable 7. Cumulative Incidence of Meeting American College of Rheumatology (ACR) Fibromyalgia Criteria Threshold Scores and Full ACR Fibromyalgia Criteria Among Consecutive Patients with PANS for Different Time Points by Age (N=204)**

| **At time (age in years)** | **Time to ACR fibromyalgia criteria threshold scores among 204 patients with PANS** | | | | **Time to full ACR fibromyalgia criteria among 204 patients with PANS** | | | |
| --- | --- | --- | --- | --- | --- | --- | --- | --- |
|  | **No. at risk** | **No. censored** | **No. events** | **Cumulative incidence, % (95% CI)** | **No. at risk** | **No. censored** | **No. events** | **Cumulative incidence, % (95% CI)** |
| 4 | 204 | 0 | 0 | 0.0 (0.0–0.0) | 204 | 0 | 0 | 0.0 (0.0–0.0) |
| 5 | 203 | 1 | 0 | 0.0 (0.0–0.0) | 203 | 1 | 0 | 0.0 (0.0–0.0) |
| 6 | 202 | 1 | 1 | 0.5 (0.1–3.4) | 203 | 1 | 0 | 0.0 (0.0–0.0) |
| 7 | 195 | 5 | 4 | 2.0 (0.7–5.2) | 199 | 5 | 0 | 0.0 (0.0–0.0) |
| 8 | 184 | 12 | 8 | 4.0 (2.0–7.9) | 190 | 13 | 1 | 0.5 (0.1–3.6) |
| 9 | 171 | 17 | 16 | 8.3 (5.2–13.2) | 182 | 20 | 2 | 1.1 (0.3–4.1) |
| 10 | 156 | 28 | 20 | 10.5 (6.9–15.8) | 169 | 32 | 3 | 1.6 (0.5–4.9) |
| 11 | 142 | 34 | 28 | 15.2 (10.7–21.2) | 160 | 41 | 3 | 1.6 (0.5–4.9) |
| 12 | 113 | 50 | 41 | 23.4 (17.7–30.5) | 138 | 59 | 7 | 4.2 (2.0–8.6) |
| 13 | 88 | 59 | 57 | 34.8 (27.9–42.8) | 125 | 69 | 10 | 6.4 (3.5–11.6) |
| 14 | 78 | 66 | 60 | 37.0 (30.0–45.2) | 110 | 81 | 13 | 8.7 (5.1–14.6) |
| 15 | 70 | 70 | 64 | 40.3 (33.0–48.7) | 97 | 88 | 19 | 13.9 (9.0–21.1) |
| 16 | 55 | 77 | 72 | 47.5 (39.5–56.1) | 82 | 100 | 22 | 16.6 (11.2–24.4) |
| 17 | 50 | 81 | 73 | 48.5 (40.4–57.2) | 68 | 111 | 25 | 19.9 (13.8–28.4) |
| 18 | 22 | 99 | 83 | 60.8 (51.6–70.0) | 41 | 138 | 25 | 19.9 (13.8–28.4) |
| 19 | 18 | 102 | 84 | 62.7 (53.2–72.2) | 36 | 142 | 26 | 22.1 (15.1–31.6) |
| 20 | 16 | 104 | 84 | 62.7 (53.2–72.2) | 28 | 149 | 27 | 24.4 (16.6–34.9) |
| 21 | 11 | 109 | 84 | 62.7 (53.2–72.2) | 18 | 158 | 28 | 27.1 (18.4–38.8) |
| 22 | 6 | 113 | 85 | 67.4 (55.4–78.9) | 9 | 167 | 28 | 27.1 (18.4–38.8) |
| 23 | 4 | 115 | 85 | 67.4 (55.4–78.9) | 6 | 170 | 28 | 27.1 (18.4–38.8) |
| 24 | 1 | 118 | 85 | 67.4 (55.4–78.9) | 3 | 173 | 28 | 27.1 (18.4–38.8) |

*Abbreviations*: American College of Rheumatology (ACR); Pediatric acute-onset neuropsychiatric syndrome (PANS); Confidence interval (CI).

**eAppendix 6. Pain Syndrome Documentation**

We observed discrepancies between questionnaire-based ACR fibromyalgia classification and EMR-documented pain syndromes:

A) Patients who met full ACR fibromyalgia criteria did not have a corresponding pain diagnosis (n=5), possibly reflecting the reluctance by providers to document a pain syndrome.
B) Patients with clinically documented pain syndromes who never met ACR fibromyalgia criteria threshold scores (n=8). Review of these eight cases identified several contributing factors. During the first year of the IBH Clinic, WPI and SSS questionnaires were not consistently collected, so we were unable to capture symptom severities from two patients with documented pain syndromes observed in this year. Two patients received pain diagnoses specific to one body region (CRPS and functional abdominal pain) thus these patients did not have a sufficient WPI score to reach the ACR fibromyalgia threshold values. The remaining four patients were ages 4-7; we have observed that young children have difficulty accurately communicating pain and fatigue symptoms to their caregivers who complete questionnaires but the fibromyalgia exam (myofascial exam) which is conducted on all of our patients, may elicit pain and further enquiry into pain that leads to a diagnosis of pain syndrome even when not recognized or reported by the caregiver on our structured questionnaire. These findings underscore the limitations of relying solely on questionnaire-based classification, especially in pediatric populations

**eFigure 3. Longitudinal Trajectories of Widespread Pain Index (WPI) Scores in Consecutive Patients with PANS (N=204)**

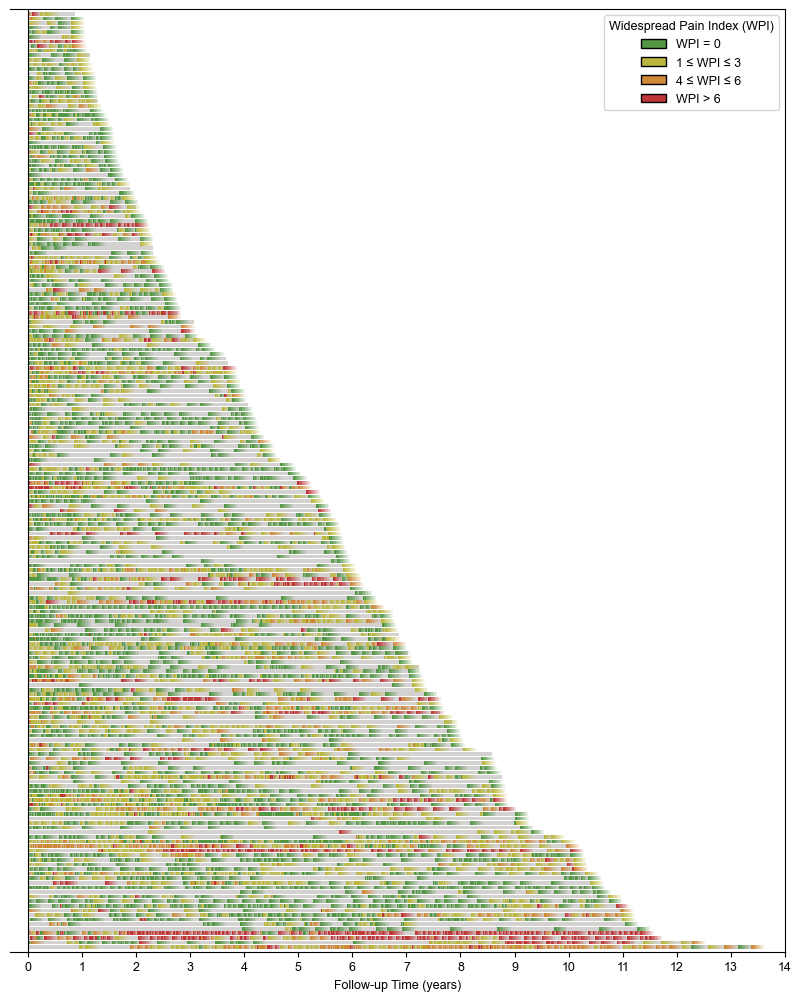

Swimmer plot depicting 204 patient trajectories over the course of follow-up by the IBH Clinic. Colored segments (green, yellow, orange) denote WPI scores collected at patient visits. Per-visit events are marked by the start of a color, followed by a fading line with length 90 days unless interrupted by a subsequent visit. Patients are ordered by duration of follow-up.

*Abbreviations*: Pediatric acute-onset neuropsychiatric syndrome (PANS); Widespread Pain Index (WPI); Immune Behavioral Health (IBH).

**eFigure 4. Longitudinal Trajectories of Waking Unrefreshed in Consecutive Patients with PANS (N=204), Part 1 of 3**

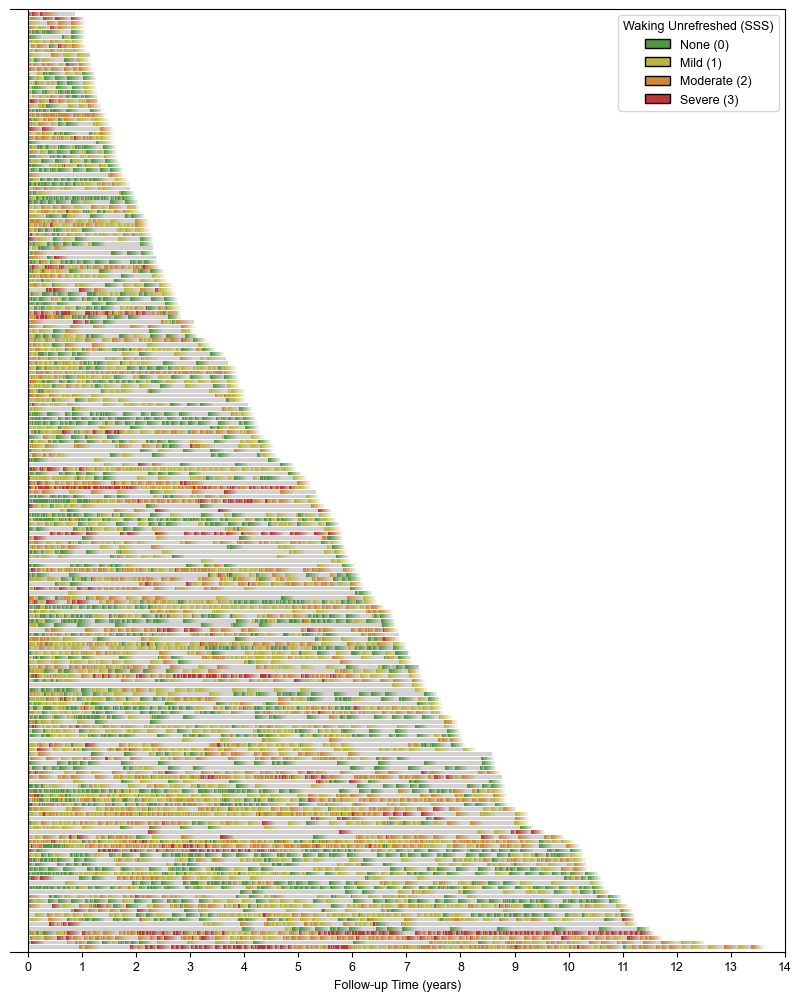

Swimmer plot depicting 204 patient trajectories over the course of follow-up by the IBH Clinic. Colored segments (green, yellow, orange, red) denote SSS (waking unrefreshed) scores collected at patient visits. Per-visit events are marked by the start of a color, followed by a fading line with length 90 days unless interrupted by a subsequent visit. Patients are ordered by duration of follow-up.

*Abbreviations*: Pediatric acute-onset neuropsychiatric syndrome (PANS); Symptom Severity Scale (SSS); Immune Behavioral Health (IBH).

**eFigure 5. Longitudinal Trajectories of Daytime Fatigue in Consecutive Patients with PANS (N=204), Part 2 of 3**

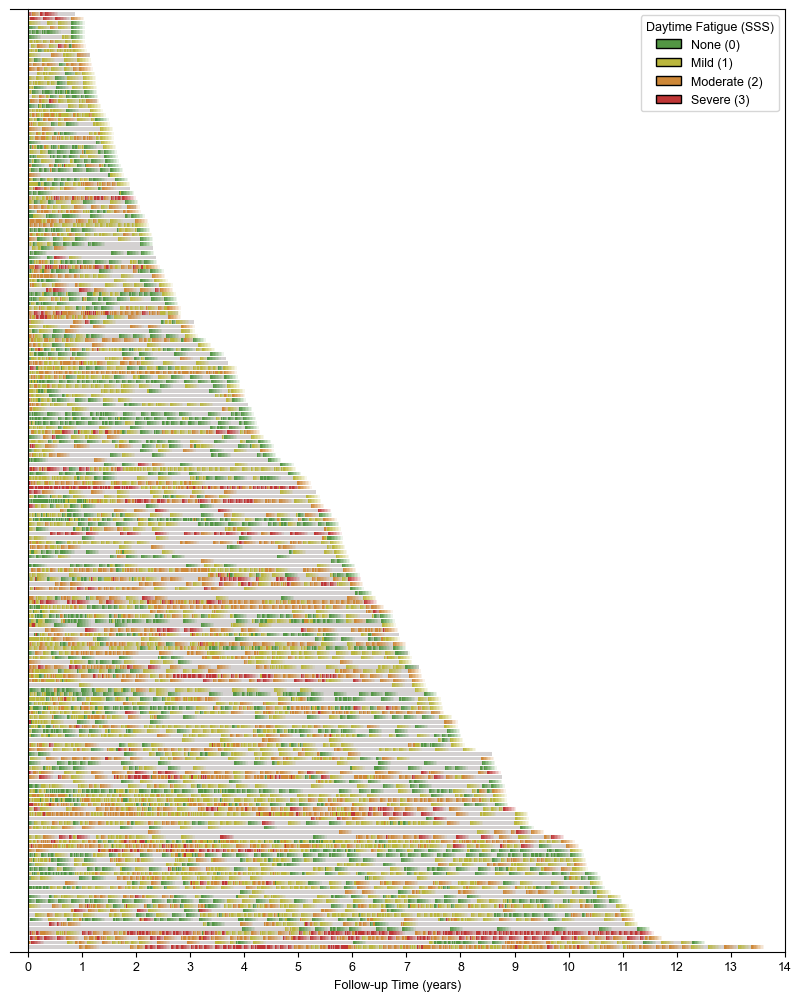

Swimmer plot depicting 204 patient trajectories over the course of follow-up by the IBH Clinic. Colored segments (green, yellow, orange, red) denote SSS (daytime fatigue) scores collected at patient visits. Per-visit events are marked by the start of a color, followed by a fading line with length 90 days unless interrupted by a subsequent visit. Patients are ordered by duration of follow-up.

*Abbreviations*: Pediatric acute-onset neuropsychiatric syndrome (PANS); Symptom Severity Scale (SSS); Immune Behavioral Health (IBH).

**eFigure 6. Longitudinal Trajectories of Cognitive Symptoms (“Brain Fog”) in Consecutive Patients with PANS (N=204), Part 3 of 3**

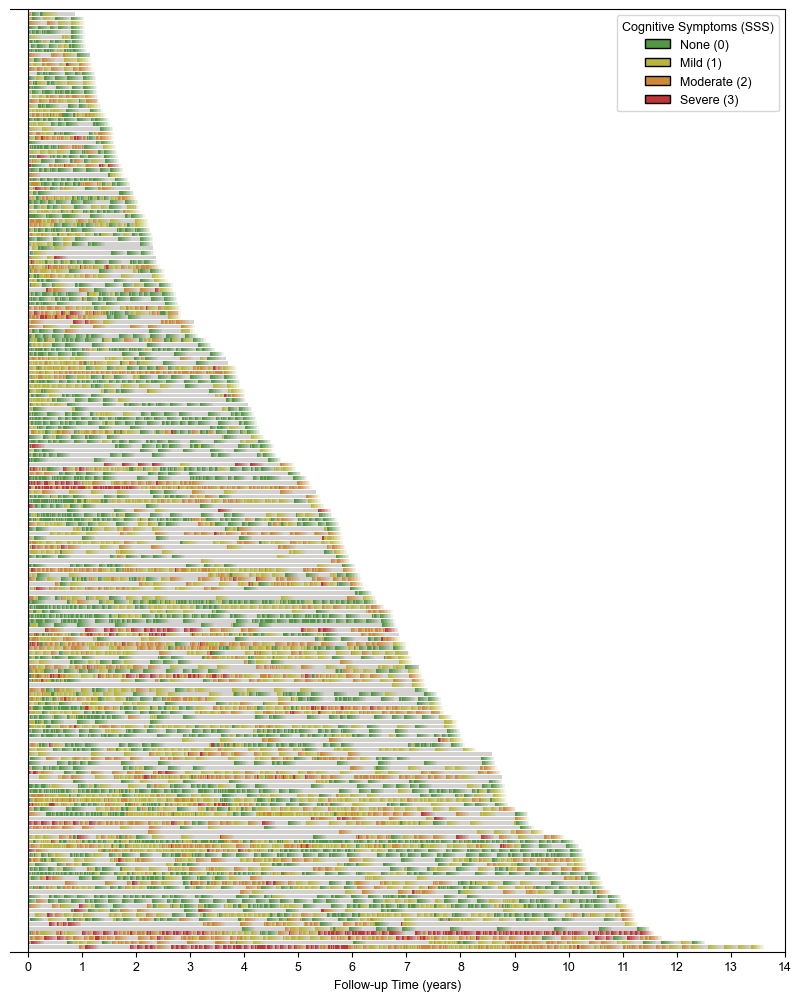

Swimmer plot depicting 204 patient trajectories over the course of follow-up by the IBH Clinic. Colored segments (green, yellow, orange, red) denote SSS (cognitive symptoms [“brain fog”]) scores collected at patient visits. Per-visit events are marked by the start of a color, followed by a fading line with length 90 days unless interrupted by a subsequent visit. Patients are ordered by duration of follow-up.

*Abbreviations*: Pediatric acute-onset neuropsychiatric syndrome (PANS); Symptom Severity Scale (SSS); Immune Behavioral Health (IBH).

**eTable 8. Exploratory Univariable Logistic Regression of Candidate Baseline Covariates**

|  | **Never Met ACR Fibromyalgia Criteria Threshold Scores (WPI and SSS)^a^** | **Met ACR Fibromyalgia Criteria Threshold Scores (WPI and SSS)^b^**  *With or without progression to full ACR fibromyalgia* | **Logistic Regression^c^** | |
| --- | --- | --- | --- | --- |
| **No.** | **104** | **64** |  |  |
| **Covariates** | **No. (%) or median [IQR]** | **No. (%) or median [IQR]** | **OR (95% CI)** | **p-value** |
| Sex (male) | 54 (51.9) | 36 (56.2) | 1.19 (0.64, 2.23) | 0.59 |
| Age at clinical presentation, years | 9.4 [6.4, 12.0] | 10.9 [9.0, 12.6] | 1.13 (1.03, 1.25) | 0.01 |
| Ever persistent at clinic intake | 5 (4.8) | 6 (9.4) | 2.05 (0.60, 7.01) | 0.25 |
| WPI at intake | 0.0 [0.0, 2.0] | 2.0 [1.0, 5.2] | 1.57 (1.31, 1.87) | <0.005 |
| SSS at intake | 4.0 [2.0, 6.0] | 7.0 [5.0, 9.0] | 1.64 (1.38, 1.94) | <0.005 |
| Sleep disturbance at intake | 50 (48.1) | 40 (62.5) | 1.80 (0.95, 3.40) | 0.07 |

^a^ ACR fibromyalgia criteria threshold scores (WPI ≥ 7 and SSS ≥ 5 OR WPI ≥ 4 and SSS ≥ 9) are derived from the ACR FSQ and based on 2016 ACR fibromyalgia criteria (**eAppendix 3**).

^b^To meet full ACR fibromyalgia criteria, patient must meet aforementioned threshold scores and have pain in 4 of 5 body regions (using the WPI) and pain must persist for 3 or more months.

^c^Univariable model. Results for each row are calculated independently of other covariates.

*Abbreviations*: American College of Rheumatology (ACR); Widespread Pain Index (WPI); Symptom Severity Scale (SSS); interquartile range (IQR); odds ratio (OR); confidence interval (CI); Fibromyalgia Survey Questionnaire (FSQ).

**eTable 9. Correlations and Variance Inflation Factors Among Covariates Included in Exploratory Cox Proportional Hazards Model**

|  | **Correlations** | | | | | | **Variance Inflation Factor** |
| --- | --- | --- | --- | --- | --- | --- | --- |
| **Covariates** | Sex (male) | Age at clinical presentation, years | Ever persistent at clinic intake | WPI at intake | SSS at intake | Sleep disturbance at intake |  |
| Sex (male) | - | 0.14 | 0.05 | -0.14 | 0.06 | -0.08 | 1.06 |
| Age at clinical presentation, years | 0.14 | - | 0.11 | 0.1 | 0.37 | -0.03 | 1.20 |
| Ever persistent at clinic intake | 0.05 | 0.11 | - | -0.01 | 0.1 | 0.05 | 1.02 |
| WPI at intake | -0.14 | 0.1 | -0.01 | - | 0.43 | 0.17 | 1.29 |
| SSS at intake | 0.06 | 0.37 | 0.1 | 0.43 | - | 0.14 | 1.45 |
| Sleep disturbance at intake | -0.08 | -0.03 | 0.05 | 0.17 | 0.14 | - | 1.05 |

*Abbreviations*: American College of Rheumatology (ACR); Widespread Pain Index (WPI); Symptom Severity Scale (SSS).

All supplemental references are also cited in the main text.
